# Implicating genes, pleiotropy and sexual dimorphism at blood lipid loci through multi-ancestry meta-analysis

**DOI:** 10.1101/2021.12.15.21267852

**Authors:** Stavroula Kanoni, Sarah E Graham, Yuxuan Wang, Ida Surakka, Shweta Ramdas, Xiang Zhu, Shoa L Clarke, Konain Fatima Bhatti, Sailaja Vedantam, Thomas W Winkler, Adam E Locke, Eirini Marouli, Greg JM Zajac, Kuan-Han H Wu, Ioanna Ntalla, Qin Hui, Derek Klarin, Austin T Hilliard, Zeyuan Wang, Chao Xue, Gudmar Thorleifsson, Anna Helgadottir, Daniel F Gudbjartsson, Hilma Holm, Isleifur Olafsson, Mi Yeong Hwang, Sohee Han, Masato Akiyama, Saori Sakaue, Chikashi Terao, Masahiro Kanai, Wei Zhou, Ben M Brumpton, Humaira Rasheed, Aki S Havulinna, Yogasudha Veturi, Jennifer Allen Pacheco, Elisabeth A Rosenthal, Todd Lingren, QiPing Feng, Iftikhar J. Kullo, Akira Narita, Jun Takayama, Hilary C Martin, Karen A Hunt, Bhavi Trivedi, Jeffrey Haessler, Franco Giulianini, Yuki Bradford, Jason E Miller, Archie Campbell, Kuang Lin, Iona Y Millwood, Asif Rasheed, George Hindy, Jessica D Faul, Wei Zhao, David R Weir, Constance Turman, Hongyan Huang, Mariaelisa Graff, Ananyo Choudhury, Dhriti Sengupta, Anubha Mahajan, Michael R Brown, Weihua Zhang, Ketian Yu, Ellen M Schmidt, Anita Pandit, Stefan Gustafsson, Xianyong Yin, Jian’an Luan, Jing-Hua Zhao, Fumihiko Matsuda, Hye-Mi Jang, Kyungheon Yoon, Carolina Medina-Gomez, Achilleas Pitsillides, Jouke Jan Hottenga, Andrew R Wood, Yingji Ji, Zishan Gao, Simon Haworth, Ruth E Mitchell, Jin Fang Chai, Mette Aadahl, Anne A Bjerregaard, Jie Yao, Ani Manichaikul, Chii-Min Hwu, Yi-Jen Hung, Helen R Warren, Julia Ramirez, Jette Bork-Jensen, Line L Kårhus, Anuj Goel, Maria Sabater-Lleal, Raymond Noordam, Pala Mauro, Floris Matteo, Aaron F McDaid, Pedro Marques-Vidal, Matthias Wielscher, Stella Trompet, Naveed Sattar, Line T Møllehave, Matthias Munz, Lingyao Zeng, Jianfeng Huang, Bin Yang, Alaitz Poveda, Azra Kurbasic, Claudia Lamina, Lukas Forer, Markus Scholz, Tessel E. Galesloot, Jonathan P. Bradfield, Sanni E Ruotsalainen, E Warwick Daw, Joseph M Zmuda, Jonathan S Mitchell, Christian Fuchsberger, Henry Christensen, Jennifer A Brody, Miguel Vazquez-Moreno, Mary F Feitosa, Mary K Wojczynski, Zhe Wang, Michael Preuss, Massimo Mangino, Paraskevi Christofidou, Niek Verweij, Jan W Benjamins, Jorgen Engmann, Noah L. Tsao, Anurag Verma, Roderick C Slieker, Ken Sin Lo, Nuno R Zilhao, Phuong Le, Marcus E Kleber, Graciela E Delgado, Shaofeng Huo, Daisuke D Ikeda, Hiroyuki Iha, Jian Yang, Jun Liu, Ayşe Demirkan, Hampton L Leonard, Jonathan Marten, Mirjam Frank, Börge Schmidt, Laura J Smyth, Marisa Cañadas-Garre, Chaolong Wang, Masahiro Nakatochi, Andrew Wong, Nina Hutri-Kähönen, Xueling Sim, Rui Xia, Alicia Huerta-Chagoya, Juan Carlos Fernandez-Lopez, Valeriya Lyssenko, Suraj S Nongmaithem, Swati Bayyana, Heather M Stringham, Marguerite R Irvin, Christopher Oldmeadow, Han-Na Kim, Seungho Ryu, Paul RHJ Timmers, Liubov Arbeeva, Rajkumar Dorajoo, Leslie A Lange, Gauri Prasad, Laura Lorés-Motta, Marc Pauper, Jirong Long, Xiaohui Li, Elizabeth Theusch, Fumihiko Takeuchi, Cassandra N Spracklen, Anu Loukola, Sailalitha Bollepalli, Sophie C Warner, Ya Xing Wang, Wen B. Wei, Teresa Nutile, Daniela Ruggiero, Yun Ju Sung, Shufeng Chen, Fangchao Liu, Jingyun Yang, Katherine A Kentistou, Bernhard Banas, Giuseppe Giovanni Nardone, Karina Meidtner, Lawrence F Bielak, Jennifer A Smith, Prashantha Hebbar, Aliki-Eleni Farmaki, Edith Hofer, Maoxuan Lin, Maria Pina Concas, Simona Vaccargiu, Peter J van der Most, Niina Pitkänen, Brian E Cade, Sander W. van der Laan, Kumaraswamy Naidu Chitrala, Stefan Weiss, Amy R Bentley, Ayo P Doumatey, Adebowale A Adeyemo, Jong Young Lee, Eva RB Petersen, Aneta A Nielsen, Hyeok Sun Choi, Maria Nethander, Sandra Freitag-Wolf, Lorraine Southam, Nigel W Rayner, Carol A Wang, Shih-Yi Lin, Jun-Sing Wang, Christian Couture, Leo-Pekka Lyytikäinen, Kjell Nikus, Gabriel Cuellar-Partida, Henrik Vestergaard, Bertha Hidalgo, Olga Giannakopoulou, Qiuyin Cai, Morgan O Obura, Jessica van Setten, Jingjing Liang, Hua Tang, Natalie Terzikhan, Jae Hun Shin, Rebecca D Jackson, Alexander P Reiner, Lisa Warsinger Martin, Zhengming Chen, Liming Li, Takahisa Kawaguchi, Joachim Thiery, Joshua C Bis, Lenore J Launer, Huaixing Li, Mike A Nalls, Olli T Raitakari, Sahoko Ichihara, Sarah H Wild, Christopher P Nelson, Harry Campbell, Susanne Jäger, Toru Nabika, Fahd Al-Mulla, Harri Niinikoski, Peter S Braund, Ivana Kolcic, Peter Kovacs, Tota Giardoglou, Tomohiro Katsuya, Dominique de Kleijn, Gert J. de Borst, Eung Kweon Kim, Hieab H.H. Adams, M. Arfan Ikram, Xiaofeng Zhu, Folkert W Asselbergs, Adriaan O Kraaijeveld, Joline WJ Beulens, Xiao-Ou Shu, Loukianos S Rallidis, Oluf Pedersen, Torben Hansen, Paul Mitchell, Alex W Hewitt, Mika Kähönen, Louis Pérusse, Claude Bouchard, Anke Tönjes, Yii-Der Ida Chen, Craig E Pennell, Trevor A Mori, Wolfgang Lieb, Andre Franke, Claes Ohlsson, Dan Mellström, Yoon Shin Cho, Hyejin Lee, Jian-Min Yuan, Woon-Puay Koh, Sang Youl Rhee, Jeong-Taek Woo, Iris M Heid, Klaus J Stark, Martina E Zimmermann, Henry Völzke, Georg Homuth, Michele K Evans, Alan B Zonderman, Ozren Polasek, Gerard Pasterkamp, Imo E Hoefer, Susan Redline, Katja Pahkala, Albertine J Oldehinkel, Harold Snieder, Ginevra Biino, Reinhold Schmidt, Helena Schmidt, Stefania Bandinelli, George Dedoussis, Thangavel Alphonse Thanaraj, Sharon LR Kardia, A Peyser, Norihiro Kato, Matthias B Schulze, Giorgia Girotto, Carsten A Böger, Bettina Jung, Peter K Joshi, David A Bennett, Philip L De Jager, Xiangfeng Lu, Vasiliki Mamakou, Morris Brown, Mark J Caulfield, Patricia B Munroe, Xiuqing Guo, Marina Ciullo, Jost B. Jonas, Nilesh J Samani, Jaakko Kaprio, Päivi Pajukanta, Teresa Tusié-Luna, Carlos A Aguilar-Salinas, Linda S Adair, Sonny Augustin Bechayda, H. Janaka de Silva, Ananda R Wickremasinghe, Ronald M Krauss, Jer-Yuarn Wu, Wei Zheng, Anneke I den Hollander, Dwaipayan Bharadwaj, Adolfo Correa, James G Wilson, Lars Lind, Chew-Kiat Heng, Amanda E Nelson, Yvonne M Golightly, James F Wilson, Brenda Penninx, Hyung-Lae Kim, John Attia, Rodney J Scott, D C Rao, Donna K Arnett, Mark Walker, Heikki A Koistinen, Giriraj R Chandak, Josep M Mercader, Teresa Tusie-Luna, Carlos Aguilar-Salinas, Clicerio Gonzalez Villalpando, Lorena Orozco, Myriam Fornage, E Shyong Tai, Rob M van Dam, Terho Lehtimäki, Nish Chaturvedi, Mitsuhiro Yokota, Jianjun Liu, Dermot F Reilly, Amy Jayne McKnight, Frank Kee, Karl-Heinz Jöckel, Mark I McCarthy, Colin NA Palmer, Veronique Vitart, Caroline Hayward, Eleanor Simonsick, Cornelia M van Duijn, Zi-Bing Jin, Jia Qu, Haretsugu Hishigaki, Xu Lin, Winfried März, Vilmundur Gudnason, Jean-Claude Tardif, Guillaume Lettre, Leen M ‘t Hart, Petra JM Elders, Daniel J Rader, Scott M Damrauer, Meena Kumari, Mika Kivimaki, Pim van der Harst, Tim D Spector, Ruth J.F. Loos, Michael A Province, Esteban J Parra, Miguel Cruz, Bruce M Psaty, Ivan Brandslund, Peter P Pramstaller, Charles N Rotimi, Kaare Christensen, Samuli Ripatti, Elisabeth Widén, Hakon Hakonarson, Struan F.A. Grant, Lambertus ALM Kiemeney, Jacqueline de Graaf, Markus Loeffler, Florian Kronenberg, Dongfeng Gu, Jeanette Erdmann, Heribert Schunkert, Paul W Franks, Allan Linneberg, J. Wouter Jukema, Amit V Khera, Minna Männikkö, Marjo-Riitta Jarvelin, Zoltan Kutalik, Cucca Francesco, Dennis O Mook-Kanamori, Ko Willems van Dijk, Hugh Watkins, David P Strachan, Niels Grarup, Peter Sever, Neil Poulter, Lee-Ming Chuang, Jerome I Rotter, Thomas M Dantoft, Fredrik Karpe, Matt J Neville, Nicholas J Timpson, Ching-Yu Cheng, Tien-Yin Wong, Chiea Chuen Khor, Hengtong Li, Charumathi Sabanayagam, Annette Peters, Christian Gieger, Andrew T Hattersley, Nancy L Pedersen, Patrik KE Magnusson, Dorret I Boomsma, Allegonda HM Willemsen, L Adrienne Cupples, Joyce B.J. van Meurs, Arfan Ikram, Mohsen Ghanbari, Penny Gordon-Larsen, Wei Huang, Young Jin Kim, Yasuharu Tabara, Nicholas J Wareham, Claudia Langenberg, Eleftheria Zeggini, Johanna Kuusisto, Markku Laakso, Erik Ingelsson, Goncalo Abecasis, John C Chambers, Jaspal S Kooner, Paul S de Vries, Alanna C Morrison, Scott Hazelhurst, Michèle Ramsay, Kari E. North, Martha Daviglus, Peter Kraft, Nicholas G Martin, John B Whitfield, Shahid Abbas, Danish Saleheen, Robin G Walters, Michael V Holmes, Corri Black, Blair H Smith, Aris Baras, Anne E Justice, Julie E Buring, Paul M Ridker, Daniel I Chasman, Charles Kooperberg, Gen Tamiya, Masayuki Yamamoto, David A van Heel, Richard C Trembath, Wei-Qi Wei, Gail P Jarvik, Bahram Namjou, M. Geoffrey Hayes, Marylyn D Ritchie, Pekka Jousilahti, Veikko Salomaa, Kristian Hveem, Bjørn Olav Åsvold, Michiaki Kubo, Yoichiro Kamatani, Yukinori Okada, Yoshinori Murakami, Bong-Jo Kim, Unnur Thorsteinsdottir, Kari Stefansson, Jifeng Zhang, Y Eugene Chen, Yuk-Lam Ho, Julie A Lynch, Daniel Rader, Philip S Tsao, Kyong-Mi Chang, Kelly Cho, Christopher J O’Donnell, John M Gaziano, Peter Wilson, Timothy M Frayling, Joel N Hirschhorn, Sekar Kathiresan, Karen L Mohlke, Million Veteran Program, Global Lipids Genetics Consortium, Yan V Sun, Andrew P Morris, Michael Boehnke, Christopher D Brown, Pradeep Natarajan, Panos Deloukas, Cristen J Willer, Themistocles L Assimes, Gina M Peloso

## Abstract

Genetic variants within nearly 1,000 loci are known to contribute to modulation of blood lipid levels. However, the biological pathways underlying these associations are frequently unknown, limiting understanding of these findings and hindering downstream translational efforts such as drug target discovery. To expand our understanding of the underlying biological pathways and mechanisms controlling blood lipid levels, we leverage a large multi-ancestry meta-analysis (N=1,654,960) of blood lipids to prioritize putative causal genes for 2,286 lipid associations by combining six gene prediction methods and assigning a confidence score. We assign, most confidently, 118 candidate causal genes and identify potential drug targets including bona-fide (*PCSK9*) and putative (*PNLIP* and *ARF6*) genes. Using phenome-wide association (PheWAS) scans, we identify relationships of genetically-predicted lipid levels to other diseases and conditions. We confirm known pleiotropic associations with cardiovascular phenotypes and determine novel associations, notably with cholelithiasis risk. We perform sex-stratified GWAS meta-analysis of lipid levels and show that 3-5% of autosomal lipid-associated loci demonstrate sex-biased effects. Finally, we report 21 novel lipid loci identified on the X chromosome. Many of the sex-biased autosomal and X chromosome lipid loci show pleiotropic associations with sex hormones, emphasizing the role of hormone regulation in lipid metabolism. Taken together, our findings provide insights into the mechanisms through which associated variants lead to altered lipid levels and potentially cardiovascular disease risk.

## INTRODUCTION

Abnormal blood lipid levels are a major cause of cardiovascular disease^1^, the leading cause of morbidity and mortality worldwide^2^. Conventional blood lipid measures, low-density lipoprotein cholesterol (LDL-C), total cholesterol (TC), triglyceride (TG), high-density lipoprotein cholesterol (HDL-C), and nonHDL-C (TC – HDL-C), are commonly used in clinical practice to identify individuals at high risk for cardiovascular events. Several treatments for reducing LDL-C, including statins, ezetimibe and PCSK9 inhibitors^3^, also reduce the risk of developing cardiovascular disease.

Genome-wide association studies (GWAS) for blood lipids have identified nearly 1,000 associated genetic loci to date^4-23^, including our recent multi-ancestry GWAS meta-analysis in 1.65M individuals^24^. The latter focused on the gains from the multi-ancestry meta-analysis relative to the single-ancestry results, in terms of number of loci, fine-mapping, and polygenic score (PGS) transferability. However, a remaining challenge is that the underlying gene and biological pathways causing the variation in lipid levels are still unknown for most of these loci. Within lipid GWAS, prior fine-mapping studies combined with functional follow-up have successfully identified causal genes with high confidence for only a handful of associated loci, including *SORT1*^25^, *TM6SF2*^12^ and *ANGPTL3*^26^, among others. Further studies with new state-of-the-art methods are needed to undertake a systematic approach to identify causal genes. Highly sophisticated methods are emerging to prioritize causal genes in well powered GWAS studies, such as the Data-driven Expression Prioritized Integration for Complex Traits^27^ (DEPICT) and the Polygenic Priority Score^28^ (PoPS), that take into account genome-wide properties of associated loci. These methods can be combined with algorithms that integrate expression data such as transcriptome-wide association studies (TWAS) and comprehensive experimental data sets such as mouse gene knock-outs. Gene sets enriched for causal genes will enhance our ability to unravel the biological pathways underlying these associations and there is growing interest in combining gene prioritization methods to provide compelling evidence for putative causal genes^29^.

In parallel, the linkage of electronic health records with genetic data in large scale population studies and patient biobanks allows for the systematic exploration of pleiotropy between traits. While blood lipid levels have a well-documented causal effect on cardiovascular disease based on genetic association studies validated by randomized controlled trials^30-32^, genetic pleiotropic associations might exist for other conditions. Unraveling such pleiotropy may yield new biological insights by revealing previously unrecognized connections between blood lipids and both cardiovascular and non-cardiovascular diseases. Phenome-wide association scans (PheWAS) adopt an agnostic approach to test for pleiotropic associations between genetic factors and a wide range of phenotypes^33^. Such knowledge may allow for the identification of lipid levels as novel diagnostic biomarkers, the repurposing of drugs, and the prevention of adverse drug events^34^.

Given empirical sex differences in blood lipid distributions, sex-specific genetic associations may yield novel biological insights. Pre-menopausal females have lower levels of LDL-C than same-age males, and HDL-C levels are higher among females of all ages compared to males^35^. Lipid levels also show a greater estimated heritability in females compared with males^36^, especially for LDL-C and TC (>1.3-fold difference). Sexual dimorphism in lipid levels may be partly explained by X chromosome variants. Evidence from human X-linked abnormalities (like Turner or Klinefelter syndromes) suggests an important role of this chromosome in lipid metabolism^37^. This is further corroborated by the lipid and atherosclerosis profiles in the Four Core Genotypes mouse model^38^, which comprises XX and XY gonadal males and XX and XY gonadal females. GWAS studies have traditionally understudied the X chromosome due to technical and analytical difficulties. A recent high coverage whole X chromosome sequencing study^39^ prioritized *CHRDL1* as a candidate causal lipid gene suggesting with larger sample sizes we may be able to discover other variation on the X chromosome associated with lipid levels.

In this study, we first identify likely causal genes at lipid loci by integrating multiple *in silico* gene prediction algorithms and experimental data sources to propose gene prioritization from GWAS using the latest Global Lipids Genetics Consortium multi-ancestry meta-analysis^24^. We then identify novel disease associations related to lipid levels through PheWAS in two large biobanks using PGSs. Finally, we perform sex-stratified meta-analysis to compare the associations between males and females to identify genetic loci with sex-specific associations and GWAS meta-analysis of the X chromosome, to better understand lipid level differences between the sexes.

## RESULTS

### Identifying functional genes in lipid associated loci

In a GWAS meta-analysis of blood lipid levels from 1.65 million individuals (**Table S1**) at 91 million genotyped and imputed genetic variants, we observed a total of 2,286 genome-wide significant index variants associated with lipid levels at 923 loci (±500 kb regions). This corresponded to 416 index variants associated with LDL-C, 539 with HDL-C, 461 with TG, 487 with TC and 383 with nonHDL-C. Uniquely, we observed 1,750 variants associated with one or more lipid levels^24^ (**Table S2**).

We employed six approaches to identify candidate functional genes for all 2,286 lipid associations and compared results for two additional approaches defined to be “gold” and “silver” standard gene sets. Our prioritization approaches include four locus-specific methods: 1) the closest gene to the index variant, 2) genes with lipid-associated protein-altering variants, 3) colocalized expression quantitative trait loci (eQTL) genes, and 4) nearby genes prioritized by transcript-wide association studies (TWAS). We also used two genome-wide approaches: 1) gene-level Polygenic Priority Score (PoPS)^28^, and 2) Data-driven Expression-Prioritized Integration for Complex Traits (DEPICT)^27^ (**Figure 1a**). We quantified how often the same gene was prioritized by multiple methods for each lipid association and determined scores that ranged from 1-6 (S1-S6) for each gene, based on the number of methods that identified the gene. For instance, a score of 6 (S6) would indicate that the same gene was identified for a variant by all six gene prioritization approaches. Since the genome-wide gene prioritization approaches can prioritize different genes for different lipid types at the same locus, we report the gene prioritization results for all 2,286 lipid-variant associations. To evaluate the performance of each prioritization method and their combined scores, we used 21 genes known to cause Mendelian dyslipidemias (**Methods**) as a gold standard set (*ABCA1, ABCG5, ANGPTL3, APOA5, APOB, APOE, CETP, CYP27A1, GPD1, GPIHBP1, LCAT, LDLR, LDLRAP1, LIPA, LIPC, LMF1, LPL, MTTP, PCSK9, SAR1B, SCARB1*), and 740 mouse knockout genes causing lipid phenotypes (**Methods**) as a silver standard set (**Table S3**).

**Figure 1.**
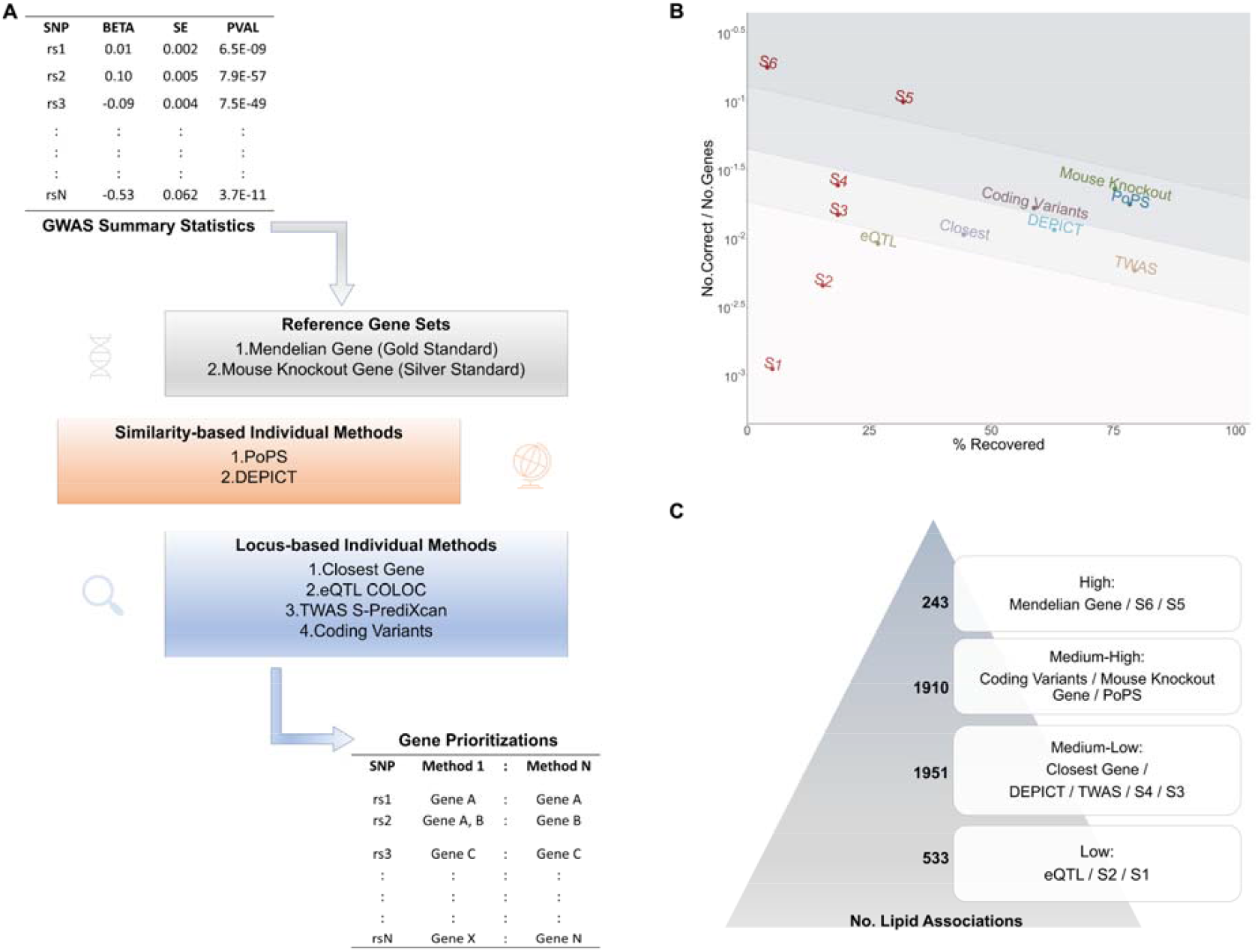
Multi-method candidate gene mapping of indexed variants associated with blood lipid levels. **A)** Schematic of gene prioritizations. We defined indexed variants within the GLGC GWAS summary statistics and performed two similarity-based methods and four locus-based methods to prioritize genes for each of the indexed variants. **B)**. Evaluation of prioritized genes. **C)**. Classification of genes into categories based on available evidence.

We examined two metrics for each gene prioritization approach: 1) the proportion of genes in the gold standard set that were prioritized, and 2) the proportion of correctly identified genes (gold standard genes) among all the genes that were nominated (**Figure 1B**). Note that out of the 2,286 lipid associations, 97 fell within 500kb of a Mendelian gene and 1,280 within 500kb of a mouse knockout gene with a lipid phenotype. Given that the closest gene is expected to be causal approximately 50% of the time^40^, we aimed to find methods that implicated a gene at a minimum of two-thirds of loci, while concurrently implicating the vast majority of gold standard genes. The proportion of gold standard genes in the gene list selected by each approach were: 79.4% by TWAS, 78.4% by PoPS, 62.9% by DEPICT, 58.8% by protein coding variants 44.3% by the closest gene and 26.8% by eQTL (**Figure S1A**). A similar rank order for the silver standard genes was observed: TWAS (42.2%), PoPS (29.8%), DEPICT (24.9%), closest gene (24.4%), coding variants (15.5%) and eQTL (12.2%) (**Figure S1B**). The TWAS method led to higher content of both Mendelian and mouse knock-out genes compared to the other methods, however, TWAS had a much smaller proportion of genes correctly prioritized, given it prioritized 3,511 genes, which was 3.5-fold greater than the other methods (∼1000 genes). We assessed lipid-relevant tissue (liver, subcutaneous and visceral adipose, whole blood and small intestine) expression QTLs (lipid eQTLs) and transcriptome-wide association (lipid TWAS) and found that the expression results from all tissues performed slightly better at recovering the reference gene sets compared with limiting to the lipid-relevant tissues (**Figure S1A** for the gold standard genes and **Figure S1B** for the silver standard genes).

We next opted to use a combined algorithm for nominating potential functional genes for each of the 2,286 lipid associations after further incorporating information from human and mouse monogenic dyslipidemias. After considering all 8 approaches (including the gold and silver sets), we assigned one of four *ad hoc* ‘confidence’ levels to the identified functional gene(s) for each index variant: high (Mendelian gene or S6 or S5), medium-high (coding variants or mouse knockout gene or PoPS), medium-low (closest gene or DEPICT or TWAS or S4 or S3) and low (eQTL or S1 or S2) (**Figure 1C**). In total, 2,373 out of 2,286 lipid associations were assigned to at least one potential causal gene; with 243 lipid associations assigned to at least one high confidence gene, 1,910 to a medium-high confidence gene, 1,951 assigned to a medium-low confidence gene and 633 index variants assigned to at least one low confidence gene (**Table S2**). When multiple genes were identified by the gene prioritization methods, we retained those with their corresponding evidence.

To assess the robustness of our prioritized genes, we applied RSS-NET^41^, a genome-wide gene prioritization approach using gene regulatory networks that was not part of our combined algorithm. Reassuringly, genes prioritized by the combined algorithm showed higher priority scores in RSS-NET than the remaining genes (maximum one-sided Wilcoxon rank sum test P-value across five lipid traits: 2.72 ×10^−250^; **Figure S2**), suggesting robustness of our prioritization approach.

We identified 118 genes as high-confidence genes among the 243 index variants, of which 97 genes were nominated by at least five methods and were not among the known Mendelian lipid genes. We performed text mining for the high-confidence genes to determine whether previous biological evidence supported these high-confidence genes as playing a role in lipid levels (**Table S4, S5**). Six out of the 97 high-confidence non-Mendelian lipid genes (*HP, STAT3, POR, PON1, FASN, CYP7A1*) have over 1000 lipid-related publications. Another 22 high-confidence non-Mendelian genes have 100-999 lipid-related publications. The remaining 63 high-confidence genes have 1-99 lipid related publications. Six high-confidence non-Mendelian genes (*ALS2CR12, BRI3, EML3, HIST1H2BF, BC1D13*, and *TMEM150A*) had no lipid-related publications retrieved by the text-mining algorithm, but we did find a subtle implication of *ALS2CR12* to lipid metabolism^42^, suggesting potential candidate genes, implicated by most of the approaches we used, for future experimental work (**Table S5**).

We performed a comprehensive look-up of all prioritized lipid genes in the Therapeutic Target Database 2020^43^ and found 9,438 drugs targeting at least one of our prioritized lipid genes (**Table S6**). Among the high-confidence and medium high-confidence prioritized genes, we identify 5,371 known drug target genes including *PCSK9* druggable as subtilisin/kexin type 9 inhibitor, *HMGCR* druggable as HMG-CoA reductase inhibitor, and *NPC1L1* druggable as Niemann-Pick C1-like protein 1 inhibitor. Additionally, *PNLIP* (pancreatic lipase) is predicted to be druggable as a pancreatic lipase inhibitor (ORLISTAT), indicating potential drug repurposing or an effect on lipids through a modification in body weight. We also identify several other potential drug targets^44^ such as *ARF6* (ADP ribosylation factor 6), *NPC1* (NPC intracellular cholesterol transporter 1), *LIPG* (lipase G) and *NR1H3* (nuclear receptor subfamily 1 group H member 3), with relevant lipid biology. *ARF6* encodes a GTP-binding protein involved in protein trafficking that regulates endocytic recycling and cytoskeleton remodeling. *NPC1* encodes an intracellular cholesterol transporter which acts in concert with NPC2 and plays an important role in the egress of cholesterol from the endosomal/lysosomal compartment. *LIPG* has phospholipase and triglyceride lipase activities and is a primary determinant of plasma HDL levels. *NR1H3* has an important role in the regulation of cholesterol homeostasis, regulating cholesterol uptake through MYLIP-dependent ubiquitination of LDLR, VLDLR and LRP8 that could be targeted as an LXR-alpha modulator.

### Effects of protective protein-altering lipid alleles on CAD, T2D and NAFLD

Coronary artery disease (CAD), type 2 diabetes (T2D), and non-alcoholic fatty liver disease (NAFLD) are typically characterized by dyslipidemias. We examined protein-altering alleles with favorable lipid profiles for their associations with CAD, T2D, and NAFLD to identify potential cardiovascular drug targets without off-target liver or diabetes effects. Of the 2,286 lipid associations, we observed 166 coding index variants. Twenty-one coding variants with a protective lipid effect also had a protective effect for CAD or T2D (p < 0.0001) and the lipid results colocalized with the CAD, or T2D results as appropriate, with a suggestive posterior probability of a shared causal variant > 0.5; **Table 1** and **S7**). Six of these twenty variants had protective effects for both CAD and T2D, while nine were protective for CAD and five were protective for T2D (**Table 1**).

**Table 1.**
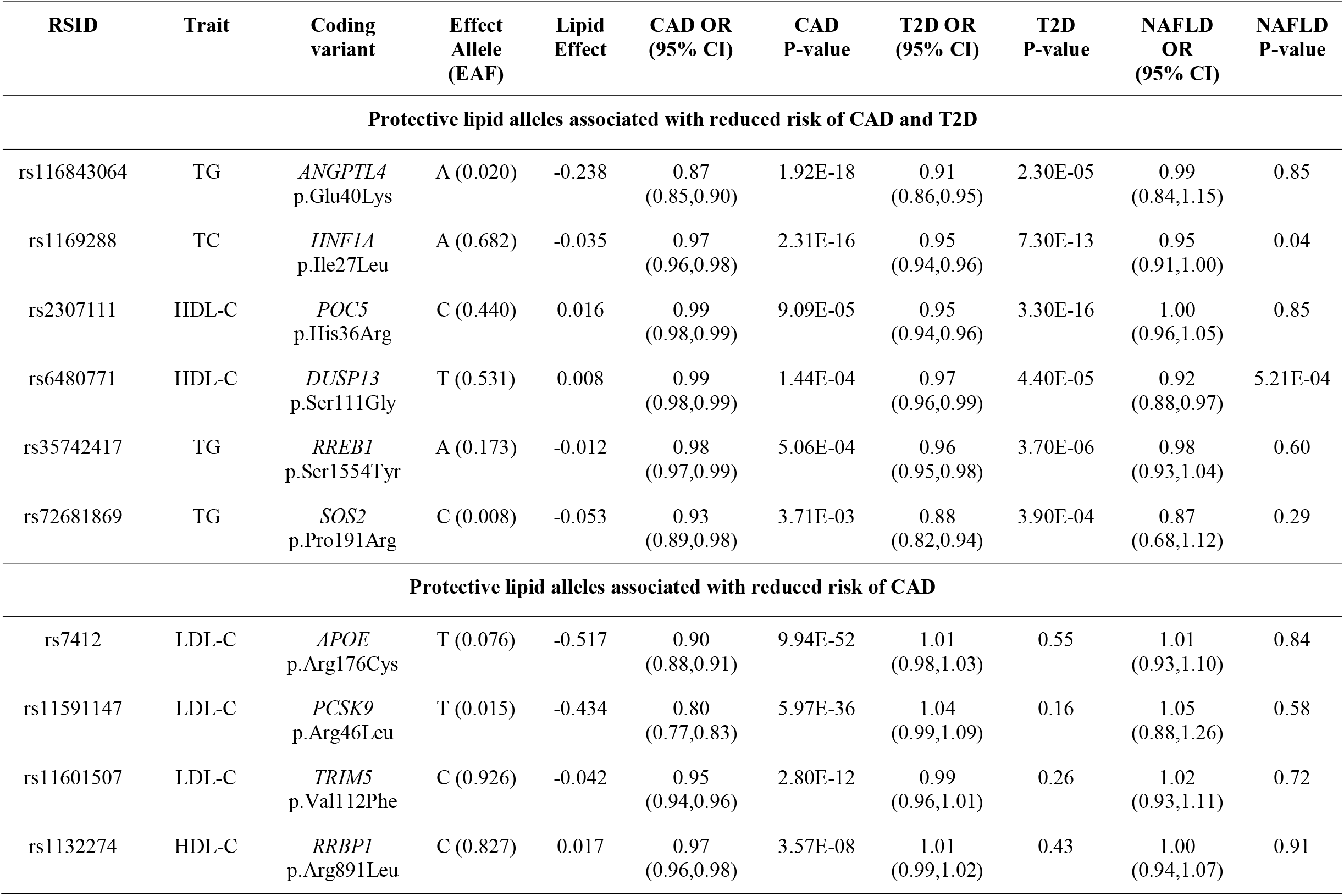
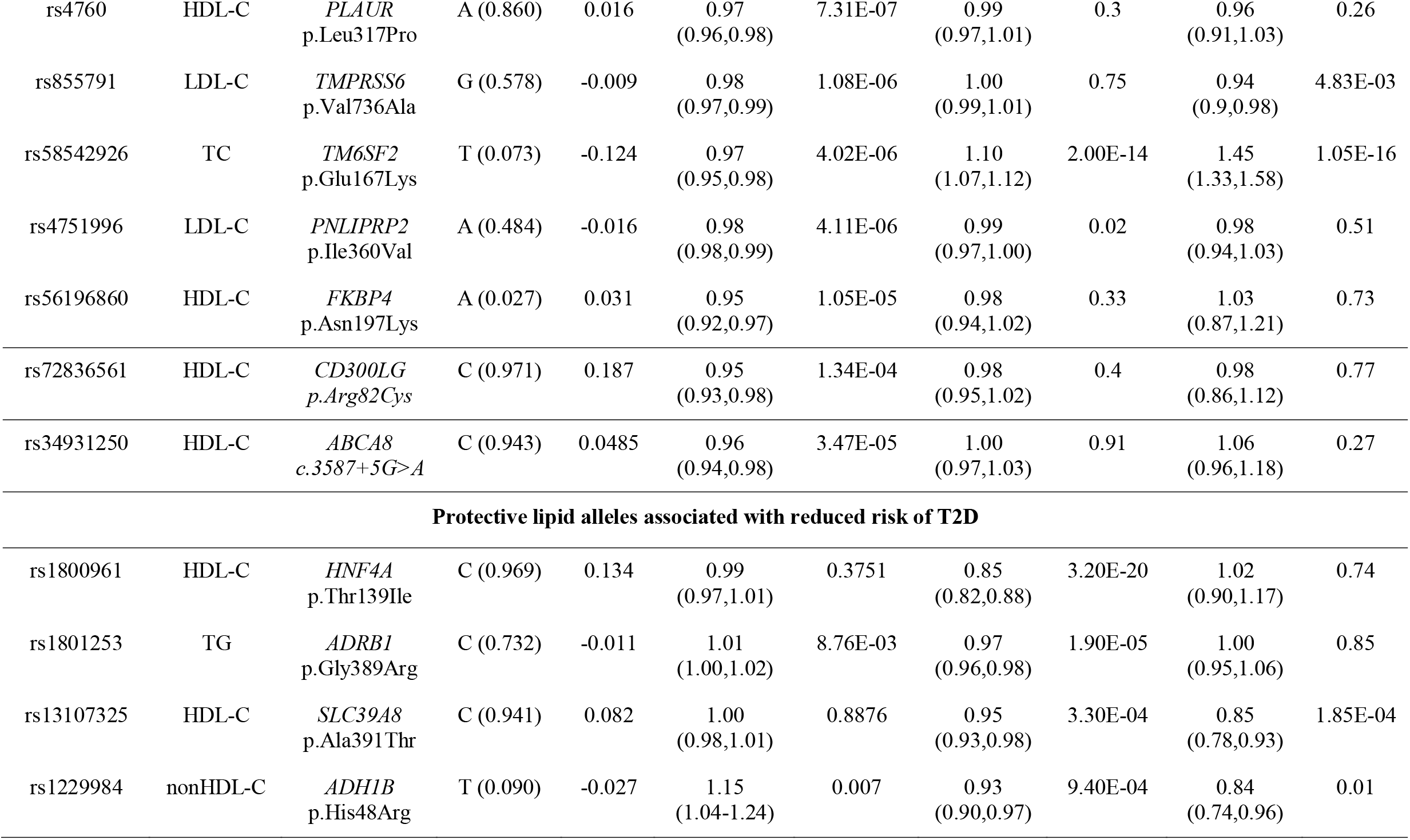
Protective lipid coding alleles associated with CAD and/or T2D.

### Polygenic scores for lipid phenotypes and phenome wide association scans

We have previously reported that a polygenic score (PGS) for LDL-C was most informative when generated from the multi-ancestry GWAS, and that the multi-ancestry PGS performed equally well in European-ancestry Americans, African-ancestry Americans, and continental Africans^24^. Using a similar approach, we generated PGS for the other four lipid traits (**Methods**).

We next performed a phenome-wide association scan (PheWAS) for the multi-ancestry lipid PGS (LDL-C PGS previously reported^24^) to identify pleiotropic effects of lipids with other traits in the European subsets of the UK Biobank and the Million Veteran Program (MVP) cohorts. We compared the effect sizes from the PheWAS analysis between UK Biobank and MVP per lipid PGS and observed a moderate correlation between the two datasets (**Figure S3**). The correlation of the PGS effects on all phenotypes between UK Biobank and MVP ranges from 0.12 for the HDL-C PGS to 0.39 for the TC PGS (**Figure S3**). In general, correlations were stronger for the ICD-10 based phecodes (r2 of 0.42-0.52) compared to the biomarkers (r2 of 0.06-0.23) (**Figure S3**), which may reflect differences in study populations due to varied environmental effects, prevalence of chronic health conditions and sex distribution. Amongst PheWAS results with p≤ 0.05 in UK Biobank, the correlation was even higher for ICD-10 based phecodes (r2 of 0.52-0.76) but remained the same for the biomarkers (r2 of 0.07-0.22).

We meta-analyzed the results from the two cohorts to increase the power of the PheWAS, by matching ICD10-mapped phecodes and biomarkers. In the combined UK Biobank-MVP PheWAS results, we detected 58 phenotypes associated with the LDL-C PGS at phenome-wide significance level (p≤6.5×10^−5^, corrected for the number of phenotypes), 165 with the HDL-C PGS, 59 with the TC PGS, 166 with the TG PGS and 78 with the non-HDL-C PGS (**Figure 2, Table S8, and Figures S4-S7**). As expected, multiple cardiovascular phenotypes related to atherosclerosis, including the expected coronary artery disease as well as aortic aneurysm and essential hypertension, were phenome-wide significantly associated with all five lipid PGSs, indicating increased risk of these diseases for individuals with genetically-predicted increased LDL-C, TG, TC or nonHDL-C or genetically-predicted decreased HDL-C. Additionally, all lipid PGSs were also significantly associated with decreased levels of direct bilirubin (**Table S8, Figures 2, S4-S7**), indicating genetically-predicted lower LDL-C increases levels of bilirubin (**Figure 2**). Correspondingly, lipid PGSs were associated with lower risk for cholelithiasis (gallstones) with the opposite direction for TG PGS, indicating that extreme lowering of LDL-C may impact rates of cholelithiasis (**Table S8, Figures 2, S4-S7**).

**Figure 2.**
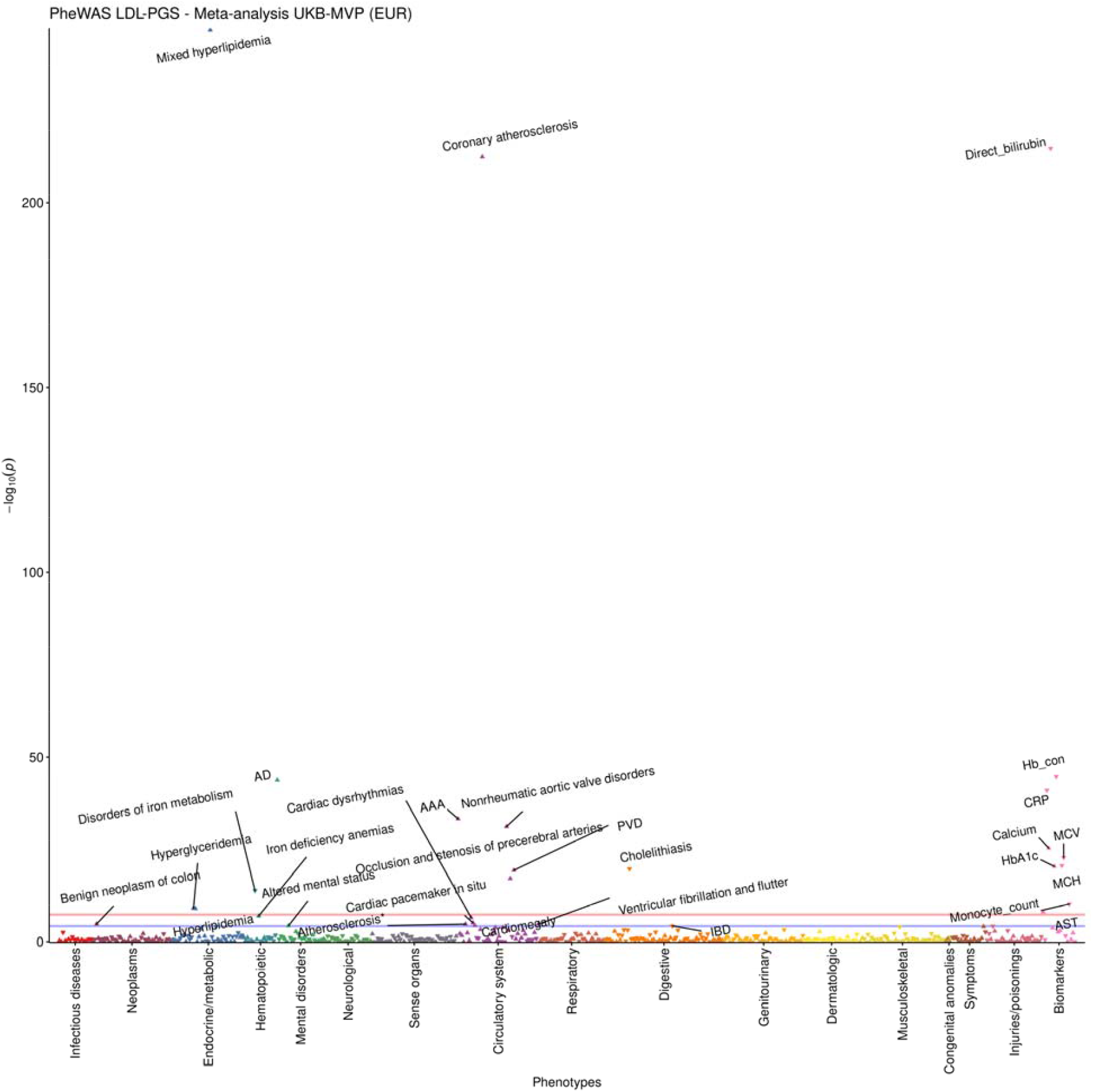
PheWAS meta-analysis results for multi-ancestry LDL-C PGS in UK Biobank and MVP. The blue horizontal line denotes phenome-wide significance (p<=6.5×10^−5^) and the red line is genome-wide significance (p<=5×10^−8^). Phenotypes have been pruned, so that the most significant one per correlated phenotype group (correlation coefficient > 0.2) is retained. Pairwise correlations were estimated with chi-square test and Cramer’s V for the dichotomous phenotypes and Pearson’s correlation for the continuous phenotypes. **AAA: Abdominal aortic aneurysm, AD: Alzheimer’s disease, AST: Aspartate aminotransferase, Atherosclerosis*: Atherosclerosis of native arteries of the extremities with intermittent claudication, Hb_con: Hemoglobin concentration, IBD: Irritable bowel disease, MCH: Mean Corpuscular Hemoglobin, MCV: Mean corpuscular volume, PVD: Peripheral vascular disease**

In the PheWAS analysis, we found that the TC and LDL-C PGS were significantly associated with increased levels of HbA1c (beta=0.101 mmol/mol per SD PGS increase, p=1.21×10^−23^ and beta=0.095 mmol/mol per SD PGS increase, p=4.37×10^−21^, respectively), while the HDL-C PGS was associated with decreased levels of HbA1c (beta=-0.257 mmol/mol per SD PGS increase, p=2.84×10^−143^) (**Table S8**). Furthermore, genetically-predicted increased LDL-C was significantly associated with decreased hemoglobin concentration (p=1.92×10^−45^, similar significant associations for all other lipid PGSs with a reverse direction of effect for TG, **Table S8**). As expected, genetically-predicted increased LDL-C and TC were both associated with increased risk for Alzheimer’s disease^45^ (OR=1.33 per SD PGS increase, p=1.74×10^−44^ and OR=1.26 per SD PGS increase, p=1.48×10^−30^, respectively; **Table S8**). Similarly, the LDL-C and TC PGSs were associated with increased aspartate aminotransferase levels (a liver enzyme), in accordance with other studies^46^. We also observed inverse associations between LDL-PGS (p=1.43×10^−14^) and TC-PGS (p=8.34×10^−14^) with the risk of iron metabolism disorders (**Table S8**).

We determined potential ancestry-specific pleiotropic effects of the multi-ancestry LDL-C PGS with coronary atherosclerosis, cholelithiasis, and HbA1c in the non-European ancestry groups in MVP. We only performed this ancestry-specific PheWAS for these selected phenotypes due to the restricted sample size. Genetically-predicted increased LDL-C was associated with decreased risk for cholelithiasis in African Americans but the opposite direction of effect was observed in Hispanic Americans or South Asians and with increased risk for coronary atherosclerosis in all ethnic groups (**Figure S8**). In African Americans, genetically-predicted increased LDL-C was associated with decreased Hb1Ac levels, in contrast to the other ancestry groups with an opposite direction of effect (**Figure S8**).

To better understand the loci that drive the association between each of the lipid PGSs and cholelithiasis and cholecystitis, we interrogated the results from the single-variant PheWAS meta-analysis in UK Biobank and MVP with all lipid multi-ancestry index variants (N=1,750 unique). We identified 22 genetic variants associated with cholelithiasis and/or cholecystitis at genome-wide significance. Genes prioritized for these index variants included genes already reported to be associated with gallstone disease^47^ (*CYP7A1, ABCG8* and *SULT2A1*), as well as additional genes (*ABCG4, SLC3A1, NAPA, ALOX5, HNF1A, RASIP1*), that may play also a role. Importantly, we found there was overlap (same index variant) between the previously published index variants for gallstone disease and our lipid index variants for these three loci (**Table S9**).

### Lipid loci show sex-specific effects

We next aimed to identify loci exhibiting differential effects on lipids between males (N=749,391) and females (N =562,410). First, we performed GWAS meta-analysis separately in each sex, excluding loci discovered in the sex-combined analysis^24^. We identified sixteen loci in females and nine loci in males that reached genome-wide significance only in one sex (p < 5×10^−8^; **Tables S10, S11** and **S12**). For example, the non-synonymous variant rs34372369 (*EPHA1*, p.Pro582Leu) is associated only in females with LDL-C, nonHDL-C, and TC (male p-values>0.05). This variant has been previously found to be linked with expression levels of the sex hormone binding globulin gene (*SHBG*) more strongly in males than females^48^, suggesting a possible reason for the difference in observed associations. However, variants may be associated in a single sex for reasons unrelated to sex differences, including due to differences in sample size between groups. We tested for a difference in effect size between males and females for these variants in an independent sex-stratified meta-analysis (from 8 independent multi-ancestry cohorts, **Table S13**), but did not find any significant differences after accounting for the number of tested variants (p-value > (0.05/16) and > (0.05/9) for females and males, respectively, **Table S14**).

Second, we tested for a difference in the male and female-specific effect sizes for each of the index variants identified from the sex-combined multi-ancestry meta-analysis. Of the 1,750 unique index variants, 64 showed a significant difference in effect size by sex for one or more traits (Bonferroni correction for the number of index variants in each trait, **Table S15**). These were evenly distributed across traits and more often had stronger effects in females than males (67%, **Figure S9)**. We tested for replication of the sex-specific differences in up to 311,120 participants from eight independent multi-ancestry cohorts not included in the original meta-analysis (**Table S13**). Fifty-four of the 64 (84%) variants had effect size differences that were directionally consistent with the original meta-analysis (**Table S16)**. Of these, 10 had significantly different effect sizes (p-value < 7.8×10^−4^, Bonferroni correction for 64 variants) and 22 were nominally significant (p-value < 0.05).

We tested whether the observed sex-differences could be caused by a higher frequency of cholesterol-lowering medications in males, potentially indicating an insufficient correction for pre-medication cholesterol levels. Among white British individuals in the UK Biobank, variants with significant sex differences had significantly higher effect size estimates on average after excluding individuals on medication (**Figure S10, Table S17**). However, of the 17 variants that exhibited a significant difference in effect size by sex in UK Biobank alone, 15 remained significant after excluding individuals taking medications. Based on this observation, the observed differences did not appear to be driven solely, or even primarily, by differences in medication use between sexes. Furthermore, none of the identified sex-specific variants were associated with sex-participation bias^49^ (**Table S18**), indicating that differential study enrollment between sexes was unlikely to be the cause of the observed sex-specific lipid associations.

Finally, we annotated each locus that showed significant sex-differences with regulatory information to identify biological mechanisms that could underlie this difference. Of the 64 lipid loci with significant sex-stratified associations, 42 showed at least a moderate colocalization (posterior probability of H4 > 0.45) with expression of 84 genes in lipid-related tissues (liver, adipose, or skeletal muscle; **Table S19**). Ten of these loci also show a sex-biased eQTL effect in at least one tissue in the direction concordant with the observed sex-specificity of the GWAS effect (**Table S19**). Amongst these ten is *CETP*, a gene with strong prior evidence for association with lipids, and *UGT2B17*^20^ (**Supplementary Note, Figure 3**). The lead variant of *UGT2B17*, rs4860987, shows a significantly stronger effect on LDL-C in males (Beta_male_=0.042, SE_male_=0.002, Beta_female_=0.016, SE_female_=0.003, p-value_difference_=4.2×10^−15^) and colocalizes with a male-specific liver eQTL associated with increased expression of *UGT2B17*. Common variants at this locus are in moderate LD (R2=0.51) with a common copy number variation (CNV), which may mediate the causal effect (**Supplementary Note**). *UGT2B17* plays a role in the metabolism of androgens^50^, including testosterone, which is consistent with the observed pleiotropic relationship of this locus with testosterone in males (**Table S19**). We note that the index variant in *UGT2B17*, rs4860987, didn’t show significant sex-specific effects in the replication cohorts, but this could be due to varying frequencies for the index variant between ancestry groups and the moderate LD to the causal CNV in the region. We observed that the combined frequency of rs4860987 across the replication studies was much lower (8%) compared with our combined frequency in the discovery (24%) due to differing proportions of ancestry groups, and along with the lower number of individuals (N=218,437), leds to a much-reduced power to replicate this sex-specific effect.

**Figure 3.**
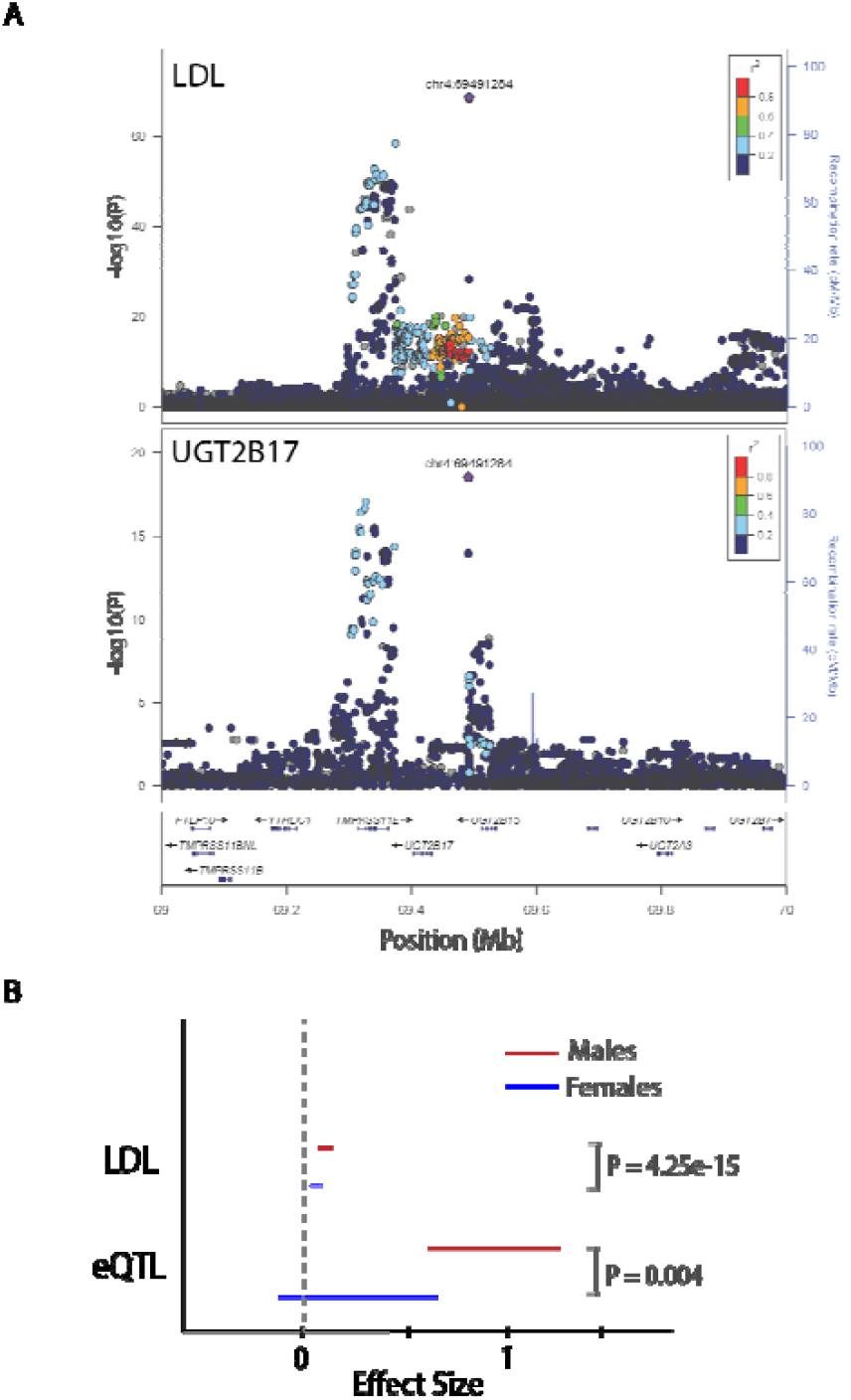
Sex-specificity at the *UGT2B17* Locus. A. The association signal for LDL-C (top panel) colocalizes with the *UGT2B17* eQTL signal in the liver (bottom panel). B The effect sizes of this variant on LDL-C and *UGT2B17* expression are both significantly higher for males (red) compared to females (blue).

### Lipid associated loci on the X chromosome

Lastly, we meta-analyzed association statistics for 3.1 million X-chromosomal variants, including PAR regions, across 1,238,180 individuals from multiple ancestry groups. We identified 28 index variants significantly associated with lipid levels (**Table S20**), of which 21 have not been previously reported^20,39,51^ (15 for HDL-C, 4 for LDL-C, 4 for TC, 5 for TG and 4 for nonHDL-C, **Table 2**). Among these 28 loci, two have index variants with a minor allele frequency (MAF) < 1% and three index variants are missense mutations (in genes *ARSL, TSPAN6* and *G6PD*), all of which are novel. We validated the identified X-chromosomal associations in up to 255,475 individuals from seven multi-ancestry cohorts (**Table S13**). Twenty index variants were at least nominally associated (p-value < 0.05), with five reaching genome-wide significance in the replication cohorts alone (p-value < 5×10^−8^, **Table S20**).

**Table 2.** Novel X-chromosome lipid associated loci

| RSID | Position in<br>chromosome<br>X (hg19) | EA/NEA | Annotation<br>(closest<br>gene) | Associated<br>Trait | EAF | N | Effect size<br>(SE)<br>from METAL | Ancestry<br>and GC<br>corrected P-value from<br>MR-MEGA | Sex<br>Difference<br>P-value |
| --- | --- | --- | --- | --- | --- | --- | --- | --- | --- |
| rs35143646 | 2856155 | T/C | Missense<br>(*ARSL*) | LDL-C | 0.6352 | 1038070 | 0.0115<br>(0.0014) | 1.83E-16 | 0.1791 |
|  |  |  |  | TC | 0.6449 | 1100310 | 0.0091<br>(0.0014) | 1.47E-14 | 0.08484 |
|  |  |  |  | nonHDL-C | 0.6822 | 712983 | 0.0112<br>(0.0017) | 2.08E-11 | 0.5294 |
| rs5934507 | 8917206 | G/A | Intergenic<br>(*FAM9B*) | HDL-C | 0.274 | 1158000 | -0.0076<br>(0.0012) | 9.99E-10 | 5.83e-4 |
| rs191084933 | 14133208 | G/A | Intergenic | HDL-C | 0.0055 | 1052630 | 0.0694 | 5.20E-10 | 0.3272 |
|  |  |  | (*GEMIN8*) |  |  |  | (0.0101) |  |  |
| rs7888119 | 16813128 | T/C | Intronic<br>(*TXLNG*) | HDL-C | 0.546 | 1158000 | 0.0114<br>(0.0012) | 1.47E-20 | 0.2212 |
| rs2230488 | 20204461 | T/G | Synonymous<br>(*RPS6KA3*) | LDL-C | 0.1772 | 1135110 | -0.0143<br>(0.0015) | 6.90E-18 | 0.9297 |
|  |  |  |  | TC | 0.1745 | 1237380 | -0.0114<br>(0.0014) | 4.68E-12 | 0.8906 |
| rs6527977 | 20322238 | A/C | Intergenic<br>(*RPS6KA3*) | nonHDL-C | 0.1602 | 789200 | -0.0147<br>(0.0020) | 3.06E-10 | 0.7000 |
| rs12012576 | 21813178 | G/A | Intergenic<br>(*SMPX*) | TG | 0.3103 | 1160340 | -0.0072<br>(0.0012) | 4.88E-8 | 0.4558 |
| rs6609434 | 46636767 | C/A | Intergenic<br>(*SLC9A7*) | HDL-C | 0.4959 | 767051 | 0.0060<br>(0.0014) | 4.25E-8 | 0.9035 |
| rs113957181 | 49848600 | A/C | Intronic<br>(*CLCN5*) | HDL-C | 0.0566 | 452268 | -0.0297<br>(0.0047) | 1.39E-9 | 0.1896 |
| rs782397956 | 53993589 | T/C | Intronic<br>(*PHF8*) | HDL-C | 0.2050 | 621342 | -0.0009<br>(0.0021) | 2.65E-15 | 0.2585 |
| rs72305711 | 55981911 | T/TTA | Intronic<br>(*RP13-188A5.1*) | HDL-C | 0.1111 | 534967 | -0.0167<br>(0.0033) | 2.22E-22 | 0.03202 |
| rs5914559 | 56139739 | G/T | Intergenic<br>(*KLF8*) | TG | 0.6697 | 738155 | -0.0131<br>(0.0016) | 4.44E-16 | 0.3224 |
| rs5964416 | 64368487 | A/C | Intergenic<br>(*ZC4H2*) | HDL-C | 0.0924 | 614283 | -0.0316<br>(0.0016) | 2.68E-11 | 0.1478 |
| rs5965342 | 66204144 | T/C | Intergenic<br>(*EDA2R*) | HDL-C | 0.2037 | 745721 | -0.0212<br>(0.0020) | 1.20E-29 | 0.02050 |
| rs505520 | 66258914 | C/A | Intergenic<br>(*EDA2R*) | TG | 0.2191 | 750415 | 0.0217<br>(0.0019) | 2.91E-30 | 0.004371 |
| rs771540123 | 67967645 | A/G | Intergenic<br>(*STARD8*) | nonHDL-C | 0.0027 | 15311 | -0.0202<br>(0.115) | 1.61E-9 | NA |
| rs5937000 | 70047788 | C/T | Intronic<br>(*TEX11*) | HDL-C | 0.4515 | 796971 | 0.0075<br>(0.0013) | 9.55E-9 | 0.02603 |
| rs5938008 | 74496225 | C/T | Intronic<br>(*UPRT*) | HDL-C | 0.9167 | 1072550 | 0.0174<br>(0.003) | 1.24E-8 | 0.7235 |
| rs1802288 | 99890204 | T/C | Missense<br>(*TSPAN6*) | LDL-C | 0.1665 | 714113 | 0.0159<br>(0.0019) | 1.42E-16 | 0.9642 |
|  |  |  |  | TC | 0.1684 | 753479 | 0.0128<br>(0.0018) | 2.49E-10 | 0.8646 |
| rs139144471 | 117829694 | G/T | Intergenic<br>(*DOCK11*) | HDL-C | 0.0876 | 1098540 | -0.0121<br>(0.0022) | 1.16E-8 | 0.1111 |
| rs6648533 | 122804678 | C/T | Intronic<br>(*THOC2*) | HDL-C | 0.299 | 1106090 | -0.0100<br>(0.0013) | 2.24E-12 | 0.4516 |
| rs5929738 | 135265287 | C/A | Intronic<br>(*FHL1*) | HDL-C | 0.4679 | 970330 | 0.0071<br>(0.0011) | 3.21E-10 | 0.4197 |
| rs5975692 | 135266089 | G/A | Intronic | TG | 0.465 | 1138740 | -0.0084 | 7.87E-12 | 0.9215 |
|  |  |  | (*FHL1*) |  |  |  | (0.0011) |  |  |
| rs2070826 | 153582198 | C/T | Intronic<br>(*FLNA*) | HDL-C | 0.1427 | 1141020 | 0.0170<br>(0.0017) | 6.65E-26 | 0.08671 |
| rs11593 | 153627145 | C/A | Intronic<br>(*RPL10*) | TG | 0.1586 | 1143360 | -0.0166<br>(0.0017) | 5.76E-21 | 0.8695 |
| rs7886627 | 153679609 | G/A | Intergenic<br>(*FAM50A*) | nonHDL-C | 0.1214 | 771706 | -0.0147<br>(0.0022) | 9.26E-9 | 0.06819 |
| rs1050828 | 153764217 | T/C | Missense<br>(*G6PD*) | LDL-C | 0.0113 | 744968 | -0.0514<br>(0.0061) | 2.54E-15 | 0.6533 |
| rs762517 | 153764734 | A/G | Intronic<br>(*G6PD*) | TC | 0.0142 | 798600 | -0.0480<br>(0.0057) | 5.26E-16 | 0.7594 |

EA=Effect allele; NEA=Non-effect allele; EAF=Effect allele frequency; N=Number of samples; SE=Standard error of the effect size; GC=Genomic control; LDL-C=Low density lipoprotein cholesterol; TC=Total cholesterol; HDL-C=High density lipoprotein cholesterol; TG=triglycerides

We additionally considered potential sex-differences for the X-chromosome variants. A missense variant in *RENBP* with MAF = 2.5% reached genome-wide significance only in males but was not significant in the sex-combined meta-analysis or in the female only analysis (p-value=4.59×10^−8^, 0.003 and 0.2, respectively). We also observe three X-chromosome loci with significant heterogeneity in effects between sexes, however, these were not significant in the replication cohorts alone, possibly due to the lower sample size (Bonferroni correction for the number of index variants in each trait, **Table S20**).

Using a PheWAS approach in the UK Biobank, we found four of the novel loci to have pleiotropic associations with body composition traits (*FAM9B* [HDL-C], *EDA2R* [HDL, TG], *TSPAN6* [LDL-C, TC], and *DOCK11* [HDL-C]), four variants with coronary atherosclerosis and ischemic heart disease, three with immune related biomarkers (*SLC9A7* [HDL-C], *CLCN5* [HDL-C], *THOC2* [HDL-C]), and two with blood clotting related biomarkers (*KLF8* [TG], *TEX11* [HDL-C]) (**Table S20**). Interestingly, two of the three sex-biased X-chromosome variants demonstrate the most significant association with testosterone of all lipid X-chromosome variants tested in the PheWAS (rs505520: beta/SE=-0.089/0.007 nmol/L per TG-increasing allele and rs5934507: beta/SE=0.237/0.006 nmol/L per HDL-increasing allele).

## DISCUSSION

In this study, we identify and prioritize likely candidate genes at lipid-associated loci discovered through a variety of approaches including multi-ancestry meta-analysis of autosomes^24^ (∼91 million variants) and the X chromosome (∼3 million variants), as well as sex-specific meta-analyses using sample sizes ranging from 1.35 to 1.65 million individuals. We previously reported a comparison of multi-ancestry vs single-ancestry lipid findings using autosomal chromosomes and identified improvements in fine-mapping of credible sets and PGS performance, with slight differences in the number of identified loci by ancestry group^24^. Here, we add X-chromosome and sex-specific discovery results and focus on genes implicated, phenotypes and diseases associated with genetically-predicted lipid levels, and prediction of drug target genes.

Our results from this effort answer three separate but related questions. We first sought to identify the optimal methods for prioritization of functional genes at GWAS loci by evaluating six methods, including their aggregation, against sets of gold and silver-standard lipid genes. Lipids are an excellent exemplar phenotype for testing of gene prioritization algorithms because of a wealth of GWAS loci (∼1000), Mendelian dyslipidemia genes (21), and mouse dyslipidemia phenotypes observed in gene knock-outs (740). After evaluating the relative performance of the six gene-prioritization methods, including the aggregation of all six methods, against Mendelian dyslipidemia genes and mouse knock-out information, we considered human Mendelian dyslipidemia genes to be most predictive, followed by mouse knock-out genes causing dyslipidemia, PoPS, protein-altering variants, DEPICT, TWAS, closest gene, and lastly eQTLs. We conclude that integrating several prioritization predictors provides higher confidence when attempting to characterize causal genes. Others have also highlighted the importance of such frameworks in different diseases^29,52,53^. However, there are notable caveats to such approaches. For example, we noticed that locus-based prioritization methods are less likely to identify multiple potential causal genes in loci with more than one independent index variant.

We then developed a scheme to classify prioritized genes at each indexed variant into 4 categories indicating our confidence with that gene being a functional lipid gene. We identify 118 unique genes with high confidence that are most likely causal as they were identified by five or more of the methods we applied (97 genes) or they were Mendelian genes (21). The vast majority of these genes had many lipid-related publications, suggesting the accuracy of our combined prioritization approach. Five high confidence genes (*BRI3, EML3, HIST1H2BF, TBC1D13*, and *TMEM150A*) had no lipid-related publications, indicating they could be truly novel or possibly were incorrectly prioritized. Functional validation of the larger pool of 2,201 medium-high confidence genes, which will require highly parallel experimental methods, will help to further optimize bioinformatics algorithms to prioritize genes.

Our prioritization approach also indicates several genes as potential novel drug targets including *PNLIP* and *ARF6. PNLIP* encodes the pancreatic lipase gene and is predicted to be druggable as a pancreatic lipase inhibitor (ORLISTAT; https://platform.opentargets.org/target/ENSG00000175535), indicating potential drug repurposing. ORLISTAT is an anti-obesity drug, with evidence for improving the lipid and glycemic profile likely secondary to weight loss^54^. The other potential novel drug target, *ARF6*, encodes the ADP-ribosylation factor 6 (Arf6), a small GTPase localized to the plasma membrane, responsible for vesicular transport and actin cytoskeleton reorganization^55^. Recently, Arf6 has been implicated with cholesterol metabolism via a phosphoinositide-based mechanism^56^ and through regulation of the ATP-binding cassette transporter 1 (ABCA1) recycling^57^. In particular, Arf6 knockdown led to ABCA1 recycling disorders that induce cholesterol accumulation in podocytes under high-glucose conditions, similar to diabetic kidney disease^57^.

We also identify twenty-one coding variants where the protective lipid allele is also protective for CAD or T2D. Among these, *PCSK9* is a well-documented drug target, not only for lipids but also for cardiovascular events^58-60^. In comparison to published studies^61^, others find a non-significant increased risk for T2D^62^ and an arguably stronger protective effect for CAD^63^, for *PCSK9* variant carriers. Our observation is consistent with the lack of excess T2D risk observed in PCSK9 inhibitor clinical trials^58-60,64^ and with strong protective effects for coronary heart disease^65^.

Our second goal was to identify diseases that may benefit from lipid-lowering as well as diseases or traits that may become problematic due to very low lipids. To accomplish this, we examined the association of genetically-predicted lipid traits (using PGS) with 773 phenotypes in 478,556 individuals. We observed, for the first time, that genetically-predicted increased LDL-C, TC and HDL-C levels, or decreased TG levels, decrease the risk of cholelithiasis. Prior epidemiological studies have not consistently reported an association between lipid levels and risk of gallstones, with some studies showing that increased levels of LDL-C, TC, and TG and decreased levels of HDL-C predispose to the risk for cholelithiasis^35,36^, but others showing no association^37,38^. The prioritized genes for the individual index lipid variants significantly associated with cholelithiasis in the PheWAS analysis include *ABCG8*, a hepatic cholesterol transporter, responsible for the efflux of cholesterol from the enterocytes to the lumen and from the hepatocytes into bile^66^. The lipid-decreasing allele of index variant in *ABCG8*, rs4245791, has been previously associated with high risk for gallstone disease^47^ and high intestinal cholesterol absorption^67^, possibly mediated by an increased expression of *ABCG8*^*68*^. These PGS-PheWAS results suggest the existence of many other cholesterol transporters like *ABCG8* that modify blood cholesterol levels perhaps in large part by facilitating an increased secretion of cholesterol into the biliary system, which in turn increases the risk of the formation of gallstones through the supersaturation of bile. We also observed that HbA1c levels were elevated among subjects with genetically-predicted increased LDL-C and TC and with genetically-predicted decreased HDL-C. Previous epidemiological studies have established associations between dyslipidemia (increased LDL-C, TC, TG and decreased HDL-C levels) and increased HbA1c levels among subjects with T2D, as well as insulin resistant subjects without diabetes^69,70^. Our observations support a strong genetic basis to these associations and are in accordance with previous studies showing shared pathways between lipid biology, T2D and HbA1c^71^, as well as pleiotropic effects of blood red cells variants with lipid levels^72^. Mendelian randomization studies have shown that hemoglobin and LDL show bidirectional inverse relationships and hemoglobin effects on LDL are also mediated through Hb1Ac, implying that genetic variation influencing erythrocytic factors could also determine lipid levels and the opposite^73^.

Lastly, we sought to expand the coverage of the genome and performed the most comprehensive GWAS of lipid levels to date by including assessment of 3 million variants on the X chromosome as well as explicitly testing for sex-specific effects across 23 chromosomes in 1.35 million individuals of diverse ancestries. We report 21 novel X-chromosome loci, including an LDL-lowering locus involving a missense variant in *G6PD*, known to cause G6PD deficiency (p.V68M)^74^. The proposed mechanism is via the inhibition of the NADPH-dependent hydroxymethylglutaryl-CoA (HMG-CoA) reductase, resulting in decreased cholesterol biosynthesis, even though the protective effect of the G6PD deficiency on cardiovascular risk is debatable^75^.

We also observed that approximately 3-5% of the genome-wide lipid index variants exhibited differential effects between sexes, which did not seem to be due to differential prevalence in the use of lipid medications or study selection bias. These findings may have important implications in the interpretation of lipid biology, the identification of novel drug targets, and possibly for more accurate prediction of blood cholesterol related conditions. For example, the *UGT2B17* locus, one of the ten sex-biased loci with corresponding sex-biased eQTL effect, is known to be implicated in androgen and drug metabolism^50^. A common CNV in the region, partially tagged by the lipid-index variant, is associated with significant variations in expression levels between ethnic groups^76^, which would explain lack of replication in the set of independent studies, and the deletion has been linked to testosterone-related decreased BMI levels^77^, as well as decreased risk for osteoporosis in men^78^.

Several of the reported sex-biased and X-chromosome loci showed significant pleiotropic effects with sex hormone levels, including testosterone and SHBG, highlighting the role of hormone regulation in lipid metabolism^79^. In particular, we observe an overall inverse effect between the X-chromosome lipid index variants and the sex-hormone levels. Observational studies have long suggested a potential influence of the sex hormones on the risk for cardiovascular risk^80^ but this hypothesis has not been consistently supported by recent Mendelian randomization studies, raising the issues of reverse causality^81,82^.

In conclusion, we leverage the power of a large multi-ancestry GWAS study to further our understanding of lipid metabolism and the impact on chronic diseases. We identify novel lipid loci on the X-chromosome and autosomal loci with evident sex-biased lipid effects. We employ a combination of prioritizing methods to identify causal genes with high confidence, an approach applicable to studying other complex traits. Our prioritized gene set can be used to point to candidate drug target genes, like *PNLIP*, without off-target adverse effects on T2D or CAD. We additionally further our understanding of lipid metabolism through a phenome-wide study that implicates a relationship between genetically-predicted low cholesterol with risk of cholelithiasis.

## METHODS

### Meta-analysis

Summary statistics for sex-combined autosomal analyses were previously published^24^. Following the same procedure, we carried out meta-analyses stratified by sex for 5 lipid traits (HDL-C, LDL-C, TG, nonHDL-C, and TC) for both the autosomes and chromosome X. The sample size for chromosome X (Total N=1,238,180; males=749,391; females =562,410) was lower than available for autosomes as not all participating biobanks submitted results for chromosome X. Quality control of summary statistics from contributing cohorts was performed using EasyQC^83^. Prior to meta-analysis, we removed variants with low imputation info scores (r^2^ < 0.3), those with minor allele count < 3, and those with Hardy-Weinberg Equilibrium p-values < 1×10^−8^. Variants on the X-chromosome were filtered using the female imputation info scores and Hardy-Weinberg Equilibrium p-values. Summary statistics were corrected by the genomic-control factor calculated from the median p-value of variants with minor allele frequency > 0.5%. For cohorts that contributed summary statistics imputed both on the Haplotype Reference Consortium (HRC) and 1000 Genomes Population v3 (1KGP3) panels, we generated a single file containing all possible variants, favoring those imputed from the HRC imputation panel due to generally higher imputation quality of these variants. Multi-ancestry meta-analysis was performed with MR-MEGA^84^ with 5 principal components and using the inverse-variance weighted method in METAL to estimate effect sizes^85^. Independent loci (**Table S21**) were defined with physical distance >500kb or genetic distance >0.25cM, whichever one would result in a larger window, followed by a conditional analysis using rareGWAMA^86^ as previously described^24^, to identify index variants that were shadows of nearby, more-significant associations. Conditional analysis for chromosome X used a female-only UK Biobank LD reference (N=21,510). In line with the analysis in the autosomes, a locus was identified as dependent if the effect size after conditioning on the most significant variant in the area was more than 1.43-times smaller than the original (95th percentile of the effect size ratios for chromosome X).

Differences in effect size between males and females were tested within each cohort using^87^:

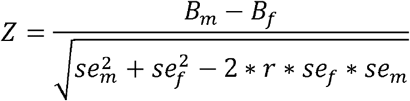

and were then meta-analyzed across studies using METAL, to account for cohort-specific ascertainment (e.g. enrichment of cases for type 2 diabetes), or demographics, such as age.

### Replication

We collected summary statistics from 8 cohorts across 6 ancestry groups, including African or African American, East Asian, European, Hispanic, Middle Eastern, and South Asian. Each cohort provided sex-stratified and X-chromosome association results for the tested traits, as available. The difference in effect sizes between males and females was calculated within each cohort as described above and then meta-analyzed across studies using METAL. X-chromosome association results were meta-analyzed using METAL with weighting by sample size.

### Gene prioritization methods

#### Closest gene

We annotated the closest gene to the lipid multi-ancestry index variants^24^ by identifying the closest gene transcript on either side (500kb) of the index variant.

#### Colocalization with GTEx eQTLs

For each of the five lipid phenotypes, we first lifted over GWAS summary statistics from the multi-ancestry meta-analysis^24^ to GrCh38 using the UCSC liftOver tool. Then, we defined a set of approximately independent windows across the genome within which co-localization with eQTLs was run. To define these, we first identified all genome-wide significant variants (P-value < 5e-08) from the meta-analysis for each lipid trait and sorted them by significance, from most significant to least. Starting with the most significant variant, we aimed to define a window defining independent genetic signals; we define a variant’s window as a region within the greater of 500kb or 0.25cM on either side of this ‘sentinel variant’. Genetic distances were defined using reference maps from HapMap 3. All other genome-wide significant variants within this window were discarded from the list of sentinel variants, and similar windows were defined for the remaining genome-wide significant variants.

We ran an eQTL colocalization using GTEx v8 eQTL summary statistics within each window for all lipid traits. For each of the 49 GTEx tissues, we first identified all genes within 1Mb of the sentinel variant, and then restricted analysis to those genes with eQTLs (‘eGenes’) in that tissue (FDR < 0.05). We used the R package ‘coloc’ (run on R version 3.4.3, coloc version 3.2.1)^88^ with default parameters to run co-localization between the GWAS signal and the eQTL signal for each of these cis-eGenes, using as input those variants in the defined window, i.e. all variants present in both datasets. A colocalization posterior probability of (PP3+PP4) > 0.8 was used to identify loci with enough colocalization power, and PP4/PP3 > 0.9 was used to define those loci that show significant colocalization^89^.

#### Transcriptome-wide association studies (TWAS)

For our transcriptome-wide association analysis (TWAS), we integrated the results of our GWAS with eQTL summary statistics from GTEx v8. The SPrediXcan software^90^ allows us to integrate these two datasets using only summary statistics from GWAS without needing individual-level genotype data. We used the multi-ancestry lipid GWAS summary statistics^24^ and harmonized them with the GTEx summary statistics. Then we performed the TWAS using the eQTL models estimated on GTEx v8 expression data. For each of the 49 GTEx tissues, we identified ‘significant genes’ those genes with P-values more significant than an FDR threshold of 0.05.

#### Genes with coding variants

We determine the coding variants within 99% credible sets and the coding variants in LD>0.8 with variants in the 99% credible sets. Additionally, we used previously established gene-based associations^91^ to determine whether rare coding variation in a gene were associated with blood lipid levels (p<0.001). We labeled a gene as having coding variants if any of these criteria were met.

#### DEPICT

We used Data-driven Expression-Prioritized Integration for Complex Traits (DEPICT, v1 beta version rel194 for 1KG imputed GWAS) to prioritize genes at our index variants, on the assumption that truly associated genes share functional annotations^27^. Index variants^24^ with p-value < 5e-8 were retained as input. We implemented the DEPICT analysis with the default settings of 500 permutations for bias adjustment and 20 replications for FDR estimation. DEPICT prioritizes genes by calculating the similarity of a given gene to genes from other associated loci across 14,461 reconstituted gene sets and estimates the nominal gene prioritization P value and the estimated false discovery rate of each gene. The prioritized genes at FDR < 0.05 were considered significant.

#### PoPS

We used the PoPS method to prioritize genes in the previously reported^24^ multi-ancestry index variants for all lipid traits. The PoPS method^28^ is a new gene prioritization method that identifies the causal genes by integrating GWAS summary statistics with gene expression, biological pathway, and predicted protein-protein interaction data. First, as part of the PoPS analysis, we used MAGMA to compute gene association statistics (z-scores) and gene-gene correlations from GWAS summary statistics and LD information from an ancestry matched reference panel (1000 Genomes). Next, PoPS performs marginal feature selection by using MAGMA to perform enrichment analysis for each gene feature separately. The model is fitted by generalized least squares (GLS), and MAGMA results are used to perform marginal feature selection, retaining only features that pass a nominal significance threshold (P < 0.05). Then PoPS, computes a joint enrichment of all selected features simultaneously in a leave one chromosome out (LOCO) framework. The gene features employed by PoPS are listed here: https://github.com/FinucaneLab/gene_features. Finally, PoPS computes polygenic priority scores for each gene by fitting a joint model for the enrichment of all selected features. The PoPS score for a gene is independent of the GWAS data on the chromosome where the gene is located. The PoPS analysis returned scores for a total of 18,383 genes per lipid trait. We only kept the top 20% genes among all 18,383 genes. We then annotated our index variants with the nearest ENSEMBL genes in a 500kb window (either side) and selected the highest PoPS score gene in the locus as the prioritized one.

We performed the PoPS analysis on our lipid-specific multi-ancestry meta-analysis results, using all populations from 1000G as the reference for the LD information in MAGMA. As a sensitivity step, we also repeated the same analysis using only the European population from 1000G as the reference. We observed high concordance in the top two PoPS prioritized genes from both reference panels. In detail, the same 2,119 genes (89%) were prioritized as the top genes from both panels, a further 203 genes were prioritized as a top gene with one panel and as the second top with the other and only 7 genes were completely mismatched between the two reference panels.

#### Overlap between methods

We integrated multiple gene prioritization methods to identify likely causal genes in the latest global lipid genetics consortium GWAS results. In total, we have implemented the 6 individual gene prioritization methods above that utilize the GWAS summary statistics from meta-analysis. Our gene prioritization methods can be placed into two broad categories, the local methods and the global methods. The local methods leverage local GWAS data by connecting the causal variants to the causal gene(s) using genomic distance, eQTLs, or protein coding variants.

More specifically, there are four local methods that have been implemented including: 1. The closest protein coding gene around the index variants based on the genomic distance, 2. eQTL colocalization using r COLOC package, 3. TWAS using S-PrediXcan, 4. Coding variants which have been identified in 99% credible sets OR in LD >0.8 with coding variants OR from gene-based tests (p <0.001) of rare coding variants. For the eQTL and TWAS we first used all the 49 GTEx tissues and then restricted to only 5 lipid specific tissues: liver, adipose subcutaneous, adipose visceral, whole blood, and small intestine. In addition, two global methods were employed: 1. DEPICT (FDR <0.05), 2. PoPS (Top 1 gene). It is reasonable to combine similarity-based methods with locus-based methods since they use two different sources of information.

We standardized the gene names across different methods using the R/geneSynonym package, a wrapper to gene synonym information in ftp://ftp.ncbi.nlm.nih.gov/gene/DATA/gene_info.gz. We also quantified how often the same gene was prioritized by multiple methods for each index variant and determined scores that ranged from 1-6 (S1-S6), based on the number of methods that prioritized the gene.

#### Monogenic genes

We annotated genes known to cause Mendelian lipid disorders based on proximity with identified GWAS loci^92,93^. GWAS index variants within +/-500kb of the transcription start and end positions from the USCS genome browser annotations were annotated as nearby known monogenic dyslipidemia genes.

#### Mouse knock-out lipid phenotypes silver set genes

Human gene symbols (9,557 unique genes) were mapped to gene identifiers (HGNC) and their corresponding mouse ortholog genes were obtained using Ensembl (www.ensembl.org). Phenotype data for single-gene knock-out mouse models were obtained from the International Mouse Phenotyping Consortium (IMPC) (www.mousephenotype.org) latest data release 12.0 (www.mousephenotype.org/data/release). The knock-out mouse models were primarily produced by IMPC but also include some models that have been reported from the relevant literature and were curated by Mouse Genome Informatics (MGI) data release 6.16 (www.informatics.jax.org). For each mouse model, reported phenotypes were grouped using the mammalian phenotype ontology hierarchy into broad categories relevant to Lipids: growth and body weight (MP:0001259), lipid homeostasis (MP:0002118), cholesterol homeostasis (MP:0005278) and lipid metabolism (MP:0013245). This resulted in mapping of human genes to significant phenotypes in animals.

For each of the multi-ancestry lipids index variant^24^, we mapped the closest gene to the knock-out mouse phenotypes and curated the set to only include mouse phenotypes strictly relating to lipid metabolism. That resulted in our silver set of 740 genes with mouse lipid phenotypes (**Table ST3**).

#### RSS-NET

We used Regression with Summary Statistics exploiting Network Topology (RSS-NET)^41^ to perform genome-wide gene prioritization analysis of five lipid traits. RSS-NET prioritized trait-associated genes by integrating GWAS summary statistics with gene regulatory networks in diverse tissues and cell types. For a given trait and network, RSS-NET produced P1, a network-specific posterior association probability for each gene^94^. To combine information from networks of multiple tissues, RSS-NET further generated a ‘meta’ P1 for each gene in each trait through Bayesian model averaging across networks. These ‘meta’ P1 values served as external gene priority scores to assess the robustness of genes prioritized by our combined algorithm (Figure S2). The RSS-NET software and related network files are available at https://github.com/suwonglab/rss-net.

#### Text mining analysis

We retrieved the whole MEDLINE/PubMed titles and abstracts as of April 30, 2021 from National Library of Medicine (https://ftp.ncbi.nlm.nih.gov/pubmed/baseline/; https://ftp.ncbi.nlm.nih.gov/pubmed/updatefiles/). We then examined whether a list of high confidence genes and any one of the lipid-related keywords (lipid, lipids, triglyceride, triglycerides, fatty acid, cholesterol, dyslipidemias, hyperlipidemia, hypercholesteremia, diabetes, type 2 diabetes, type II diabetes, heart, cardiovascular, artery, coronary, coronary artery, coronary heart, atherosclerosis, peripheral vascular, PAD, stroke) occurred in the same abstract. We counted how many lipid-related publications that have a specific gene co-occurred with at least one lipid-related keyword.

#### Drug targets mining analysis

To gain therapeutic insights from our gene prioritization results, we performed a lookup in Therapeutic Target Database (TTD) 2020^43^ (http://db.idrblab.net/ttd/). Specifically, we cross-referenced 5,710 unique lipid-associated genes prioritized by our combined algorithm (**Table S2**) with 1,563 genes corresponding to at least one drug (either under development or approved) with known clinical indication in TTD 2020. As a quality control for this lookup, we excluded all TTD entries related to drugs that were discontinued, terminated, or withdrawn from the market. The full lookup results are available in **Table S6**.

### CAD/T2D colocalization analysis with lipid traits

We used R package coloc v3.2.1^88^ to perform summary statistics-based colocalization via a Bayesian approach and test whether the 5 lipid traits share common genetic causal variants with CAD or T2D. We first defined a window of +/-100KB around each index variant^24^. Then for each window of the 10 pairs of traits, we ran colocalization with default parameters using those SNPs present in both datasets. A colocalization posterior probability of PP4 > 0.5 was used to define those loci that show significant colocalization.

### PheWAS analysis

#### Construction of lipid PGSs

We had previously developed a multi-ancestry PGS for LDL-C that was demonstrated to perform well across multiple ancestry groups^24^. In a similar manner, we also generated PGS for HDL-C, nonHDL-C, TC, and triglycerides. First, multi-ancestry meta-analysis results were generated with METAL^85^ after excluding individuals from the Michigan Genomics Initiative and the UK Biobank. The set of variants used to construct the PGS was limited to those that were well-imputed (r^2^ > 0.3) in MGI, UK Biobank, and MVP. Risk scores based on PRS-CS^95^ or pruning and thresholding with Plink^96^ across several r^2^ (0.1, 0.2), distance (250kb, 500kb), and p-value thresholds (5×10^−10^, 5×10^−9^, 5×10^−8^, 5×10^−7^, 5×10^−6^, 5×10^−5^, 5×10^−4^, 5×10^−3^, 0.05) were developed. For each trait, the single best score was selected based on the adjusted r^2^ calculated in UK Biobank of the linear model for the lipid trait with the risk score and age, sex, batch, and PC1-4 as covariates. This corresponded to PRS-CS for HDL-C and nonHDL-C and pruning and thresholding for LDL-C (r^2^=0.1, p-value=5×10^−4^, 500kb), TG (r^2^=0.1, p-value=5×10^−3^, 500kb), and TC (r^2^=0.1, p-value=5×10^−4^, 500kb). The variance explained by the risk score among UK Biobank participants was similar across traits (adjusted r^2^ of the full model - adjusted r^2^ of covariates: HDL-C=0.13; LDL-C=0.15; nonHDL-C=0.14; TC=0.14; TG=0.10) and validated the ability of the risk score to predict genetically increased lipid levels.

#### PheWAS of lipid PGSs and index lipid variants in UK Biobank and MVP

We used the European ancestry subset of individuals from UK Biobank (408,886 samples) and the European samples from MVP (69,670 samples) to perform the PheWAS analysis.

We constructed a weighted PGS for each of the lipid traits, based on the corresponding genome-wide significant multi-ancestry index variants. We used the PheWAS package in R^97^ to map ICD-10 codes from hospital records into clinically relevant phenotypes (phecodes) and to implement these association analysis, while adjusting for sex, age, 10 genetic principal components, genotyping array (for UK Biobank only) in each cohort. For the lipid-PGS PheWAS, each PGS was inverse normalized prior to analysis and lipid levels were corrected for statin use. The MVP samples used for the PheWAS analysis were not included in the GWAS meta-analysis^24^.

Similarly, we extracted all multi-ancestry autosomal index variants for all lipid traits from the same European ancestry subset of UK Biobank and MVP and performed a single-variant PheWAS association analysis per cohort. Additionally, we performed a single variant PheWAS association analysis in UK Biobank only with the sex-stratified and X-chromosome index variants from the multi-ancestry analysis.

#### Meta-analysis of MVP and UK Biobank PheWAS results

We combined, via meta-analysis, PheWAS lipid-specific PGS results for all intersecting phecodes and biomarkers between UK Biobank and MVP (Europeans only) per lipid trait. We used ICD10-based phecodes and manually-matched biomarkers to identify intersecting phenotypes between the two datasets. We restricted our meta-analysis to phenotypes that had at least 100 samples (total number for continuous traits or number of cases for binary traits) in each cohort. After the meta-analysis, we excluded phenotypes that had less than 500 combined samples (total number for continuous traits or number of cases for binary traits), to avoid reporting spurious results^98^. That resulted in a total of 773 phenotypes (739 phecodes and 34 biomarkers/measurements). We used both fixed and random effects model for the meta-analysis. We assessed heterogeneity using the p-value for Cochran’s q and set the level for significant heterogeneity at a Bonferroni threshold. We report the results from the fixed-effects model for the phenotypes with non-significant heterogeneity and the results from the random-effects model for all others. For most significant and novel findings, we performed the PheWAS analysis in the African American and Hispanic subset of the MVP study. Similarly, we meta-analysed all index-variant PheWAS results between UK Biobank and MVP and obtained results for 811 phenotypes and 1,750 lipid multi-ancestry index variants, after excluding instances with a combined sample size < 500.

### Lipid index variants with CAD, T2D and NAFLD datasets

The GWAS meta-analysis results of CAD and T2D were acquired from MVP^63^ and DIAGRAM Consortium^62^, respectively. For variant rs1229984, the CAD result is from CARDIoGRAM*Plus*C4D meta-analysis^99^, as it was not present in the MVP results. The NAFLD GWAS and meta-analysis was performed in the UK Biobank and Michigan Initiative (MGI). We determined the association of the lipid index variants with CAD, T2D, and NAFLD, and aligned the alleles across all the traits to the LDL lowering allele. We then highlighted the protective lipid coding alleles associated with CAD.

#### GWAS and Meta-analysis of NAFLD in UK Biobank and Michigan Genomics Initiative (MGI)

Individuals with NAFLD were identified using ICD-9 571.8 and ICD-10 K76.0. Individuals with hepatitis, liver cirrhosis, liver abscess, ascites, a liver transplant, hepatomegaly, jaundice, or with abnormal result of serum enzyme levels or a function study of the liver were excluded (exclusion phecodes 70.2, 70.3, 571.51, 571.6, 571.8, 571.81, 572, 573, 573.2, 573.3, 573.5, 573.7, 573.9)^100^. Analysis was performed using SAIGE v43.3^101^. Analysis in the UK Biobank included white British individuals with batch, sex, birth year, and the first 4 genetic principal components as covariates. A total of 1,122 cases and 399,900 controls were included in the analysis. Analysis in MGI included only European ancestry participants with array version, sex, birth year, and the first 4 genetic principal components as covariates. A total of 2,901 cases and 49,098 controls were analyzed. Meta-analysis was performed using METAL with weighting based on the effective sample size calculated as 4/((1/Ncases)+(1/Ncontrols)).

## Data Availability

All data produced in the present study are available upon reasonable request to the authors

## Data Availability

The GWAS meta-analysis results (including both ancestry-specific and trans-ancestry analyses) and risk score weights are available at: http://csg.sph.umich.edu/willer/public/glgc-lipids2021. The optimized trans-ancestry polygenic score weights will be deposited within the PGS Catalog (https://www.pgscatalog.org/).

## Code Availability

Scripts used for analysis and summary of results are available upon request.

## Conflicts of Interest

Ioanna Ntalla is an employee and stock owner of Gilead Sciences since August 2019. Derek Klarin accepts consulting fees from Regeneron Pharmaceuticals. All deCODE affiliated authors (Gudmar Thorleifsson, Anna Helgadottir, Daniel F Gudbjartsson, Hilma Holm, Unnur Thorsteinsdottir, Kari Stefansson) are employees of deCODE/Amgen Inc. As of January 2020, Anubha Mahajan is an employee of Genentech, and a holder of Roche stock. Markus Scholz receives funding from Pfizer Inc. for a project not related to this research. Marcus E Kleber is employed by SYNLAB MVZ Mannheim GmbH. Gabriel Cuellar-Partida contributed to this work while employed at The University of Queensland, but he is now an employee of 23andMe Inc. Mark J Caulfield is Chief Scientist for Genomics England, a UK Government company. The views expressed in this article are those of the author(s) and not necessarily those of the NHS, the NIHR, or the Department of Health. Mark I McCarthy has served on advisory panels for Pfizer, NovoNordisk and Zoe Global, has received honoraria from Merck, Pfizer, Novo Nordisk and Eli Lilly, and research funding from Abbvie, Astra Zeneca, Boehringer Ingelheim, Eli Lilly, Janssen, Merck, NovoNordisk, Pfizer, Roche, Sanofi Aventis, Servier, and Takeda. As of June 2019, Mark I McCarthy is an employee of Genentech, and a holder of Roche stock. Winfried März has received grants from Siemens Healthineers, grants and personal fees from Aegerion Pharmaceuticals, grants and personal fees from AMGEN, grants from Astrazeneca, grants and personal fees from Sanofi, grants and personal fees from Alexion Pharmaceuticals, grants and personal fees from BASF, grants and personal fees from Abbott Diagnostics, grants and personal fees from Numares AG, grants and personal fees from Berlin-Chemie, grants and personal fees from Akzea Therapeutics, grants from Bayer Vital GmbH, grants from bestbion dx GmbH, grants from Boehringer Ingelheim Pharma GmbH Co KG, grants from Immundiagnostik GmbH, grants from Merck Chemicals GmbH, grants from MSD Sharp and Dohme GmbH, grants from Novartis Pharma GmbH, grants from Olink Proteomics, other from Synlab Holding Deutschland GmbH, all outside the submitted work. Bruce M Psaty serves on the Steering Committee of the Yale Open Data Access Project funded by Johnson & Johnson. Amit V Khera has served as a consultant to Sanofi, Medicines Company, Maze Pharmaceuticals, Navitor Pharmaceuticals, Verve Therapeutics, Amgen, and Color Genomics; received speaking fees from Illumina, the Novartis Institute for Biomedical Research; received sponsored research agreements from the Novartis Institute for Biomedical Research and IBM Research, and reports a patent related to a genetic risk predictor (20190017119). Dennis O Mook-Kanamori is a part-time clinical research consultant for Metabolon, Inc. Danish Saleheen has received support from the British Heart Foundation, Pfizer, Regeneron, Genentech, and Eli Lilly pharmaceuticals. Veikko Salomaa has received honoraria for consultations from Novo Nordisk and Sanofi and has ongoing research collaboration with Bayer Ltd, all unrelated to the present study. Sekar Kathiresan is an employee of Verve Therapeutics, and holds equity in Verve Therapeutics, Maze Therapeutics, Catabasis, and San Therapeutics. He is a member of the scientific advisory boards for Regeneron Genetics Center and Corvidia Therapeutics; he has served as a consultant for Acceleron, Eli Lilly, Novartis, Merck, Novo Nordisk, Novo Ventures, Ionis, Alnylam, Aegerion, Haug Partners, Noble Insights, Leerink Partners, Bayer Healthcare, Illumina, Color Genomics, MedGenome, Quest, and Medscape; he reports patents related to a method of identifying and treating a person having a predisposition to or afflicted with cardiometabolic disease (20180010185) and a genetics risk predictor (20190017119). Cristen J Willer’s spouse is employed by Regeneron.

## Acknowledgments

GMP, PN and CW are supported by NHLBI R01HL127564. GMP and PN are supported by R01HL142711. AG acknowledge support from the Wellcome Trust (201543/B/16/Z), European Union Seventh Framework Programme FP7/2007-2013 under grant agreement no. HEALTH-F2-2013-601456 (CVGenes@Target) & the TriPartite Immunometabolism Consortium [TrIC]-Novo Nordisk Foundation’s Grant number NNF15CC0018486. JMM is supported by American Diabetes Association Innovative and Clinical Translational Award 1-19-ICTS-068. SR was supported by the Academy of Finland Center of Excellence in Complex Disease Genetics (Grant No 312062), the Finnish Foundation for Cardiovascular Research, the Sigrid Juselius Foundation and University of Helsinki HiLIFE Fellow and Grand Challenge grants. EW was supported by the Finnish innovation fund Sitra (EW) and Finska Läkaresällskapet. CNS was supported by American Heart Association Postdoctoral Fellowships 15POST24470131 and 17POST33650016. Charles N Rotimi is supported by Z01HG200362. Zhe Wang, Michael Presuss, and Rith JF Loos are supported by R01HL142302. NJT is a Wellcome Trust Investigator (202802/Z/16/Z), is the PI of the Avon Longitudinal Study of Parents and Children (MRC & WT 217065/Z/19/Z), is supported by the University of Bristol NIHR Biomedical Research Centre (BRC-1215-2001), the MRC Integrative Epidemiology Unit (MC_UU_00011) and works within the CRUK Integrative Cancer Epidemiology Programme (C18281/A19169). Ruth E Mitchell is a member of the MRC Integrative Epidemiology Unit at the University of Bristol funded by the MRC (MC_UU_00011/1). Simon Haworth is supported by the UK National Institute for Health Research Academic Clinical Fellowship. Paul S. de Vries was supported by American Heart Association grant number 18CDA34110116. Julia Ramierz acknowledges support by the People Programme of the European Union’s Seventh Framework Programme grant n° 608765 and Marie Sklodowska-Curie grant n° 786833. Maria Sabater-Lleal is supported by a Miguel Servet contract from the ISCIII Spanish Health Institute (CP17/00142) and co-financed by the European Social Fund. Jian Yang is funded by the Westlake Education Foundation. Olga Giannakopoulou has received funding from the British Heart Foundation (BHF) (FS/14/66/3129). CHARGE Consortium cohorts were supported by R01HL105756. Study specific acknowledgement are available in the **Supplementary Material**.

## Notes

### Funding Statement

A detailed funding statement is included in the supplement.

### Author Declarations

This study was approved by the IRB of the Boston University Medical Center (IRB Number: H-41599).

## REFERENCES

1. Castelli, W.P., Anderson, K., Wilson, P.W. & Levy, D. Lipids and risk of coronary heart disease. The Framingham Study. Ann Epidemiol 2, 23–8 (1992).

2. GBD 2019 Diseases and Injuries Collaborators. Global burden of 369 diseases and injuries in 204 countries and territories, 1990-2019: a systematic analysis for the Global Burden of Disease Study 2019. Lancet 396, 1204–1222 (2020).

3. Grundy, S.M. et al. 2018 AHA/ACC/AACVPR/AAPA/ABC/ACPM/ADA/AGS/APhA/ASPC/NLA/PCNA Guideline on the Management of Blood Cholesterol: A Report of the American College of Cardiology/American Heart Association Task Force on Clinical Practice Guidelines. Circulation 139, e1082–e1143 (2019).

4. Diabetes Genetics Initiative of Broad Institute of Harvard and MIT, Lund University, and Novartis Institutes of BioMedical Research. et al. Genome-wide association analysis identifies loci for type 2 diabetes and triglyceride levels. Science 316, 1331–6 (2007).

5. Kathiresan, S. et al. A genome-wide association study for blood lipid phenotypes in the Framingham Heart Study. BMC Med Genet 8 Suppl 1, S17 (2007).

6. Kathiresan, S. et al. Polymorphisms associated with cholesterol and risk of cardiovascular events. N Engl J Med 358, 1240–9 (2008).

7. Teslovich, T.M. et al. Biological, clinical and population relevance of 95 loci for blood lipids. Nature 466, 707–13 (2010).

8. Asselbergs, F.W. et al. Large-scale gene-centric meta-analysis across 32 studies identifies multiple lipid loci. Am J Hum Genet 91, 823–38 (2012).

9. Albrechtsen, A. et al. Exome sequencing-driven discovery of coding polymorphisms associated with common metabolic phenotypes. Diabetologia 56, 298–310 (2013).

10. Tachmazidou, I. et al. A rare functional cardioprotective APOC3 variant has risen in frequency in distinct population isolates. Nat Commun 4, 2872 (2013).

11. Willer, C.J. et al. Discovery and refinement of loci associated with lipid levels. Nat Genet 45, 1274–1283 (2013).

12. Holmen, O.L. et al. Systematic evaluation of coding variation identifies a candidate causal variant in TM6SF2 influencing total cholesterol and myocardial infarction risk. Nat Genet 46, 345–51 (2014).

13. Peloso, G.M. et al. Association of low-frequency and rare coding-sequence variants with blood lipids and coronary heart disease in 56,000 whites and blacks. Am J Hum Genet 94, 223–32 (2014).

14. Surakka, I. et al. The impact of low-frequency and rare variants on lipid levels. Nat Genet 47, 589–97 (2015).

15. Tang, C.S. et al. Exome-wide association analysis reveals novel coding sequence variants associated with lipid traits in Chinese. Nat Commun 6, 10206 (2015).

16. van Leeuwen, E.M. et al. Genome of The Netherlands population-specific imputations identify an ABCA6 variant associated with cholesterol levels. Nat Commun 6, 6065 (2015).

17. Iotchkova, V. et al. Discovery and refinement of genetic loci associated with cardiometabolic risk using dense imputation maps. Nat Genet 48, 1303–1312 (2016).

18. Liu, D.J. et al. Exome-wide association study of plasma lipids in >300,000 individuals. Nat Genet 49, 1758–1766 (2017).

19. Lu, X. et al. Exome chip meta-analysis identifies novel loci and East Asian-specific coding variants that contribute to lipid levels and coronary artery disease. Nat Genet 49, 1722–1730 (2017).

20. Hoffmann, T.J. et al. A large electronic-health-record-based genome-wide study of serum lipids. Nat Genet 50, 401–413 (2018).

21. Kanai, M. et al. Genetic analysis of quantitative traits in the Japanese population links cell types to complex human diseases. Nat Genet 50, 390–400 (2018).

22. Klarin, D. et al. Genetics of blood lipids among ∼300,000 multi-ethnic participants of the Million Veteran Program. Nat Genet 50, 1514–1523 (2018).

23. Spracklen, C.N. et al. Association analyses of East Asian individuals and trans-ancestry analyses with European individuals reveal new loci associated with cholesterol and triglyceride levels. Hum Mol Genet 27, 1122 (2018).

24. Graham, S.E. et al. The power of genetic diversity in genome-wide association studies of lipids. Nature (in press) (2021).

25. Musunuru, K. et al. From noncoding variant to phenotype via SORT1 at the 1p13 cholesterol locus. Nature 466, 714–9 (2010).

26. Musunuru, K. et al. Exome sequencing, ANGPTL3 mutations, and familial combined hypolipidemia. N Engl J Med 363, 2220–7 (2010).

27. Pers, T.H. et al. Biological interpretation of genome-wide association studies using predicted gene functions. Nat Commun 6, 5890 (2015).

28. Weeks, E.M. et al. Leveraging polygenic enrichments of gene features to predict genes underlying complex traits and diseases. medRxiv, 2020.09.08.20190561 (2020).

29. Stanzick, K.J. et al. Discovery and prioritization of variants and genes for kidney function in >1.2 million individuals. Nature Communications 12, 4350 (2021).

30. The Emerging Risk Factors Collaboration. et al. Major lipids, apolipoproteins, and risk of vascular disease. JAMA 302, 1993–2000 (2009).

31. Richardson, T.G. et al. Evaluating the relationship between circulating lipoprotein lipids and apolipoproteins with risk of coronary heart disease: A multivariable Mendelian randomisation analysis. PLoS Med 17, e1003062 (2020).

32. Allara, E. et al. Genetic Determinants of Lipids and Cardiovascular Disease Outcomes. Circulation: Genomic and Precision Medicine 12, e002711 (2019).

33. Veturi, Y. et al. A unified framework identifies new links between plasma lipids and diseases from electronic medical records across large-scale cohorts. Nat Genet 53, 972–981 (2021).

34. Bush, W.S., Oetjens, M.T. & Crawford, D.C. Unravelling the human genome-phenome relationship using phenome-wide association studies. Nat Rev Genet 17, 129–45 (2016).

35. Abbott, R.D. et al. Joint distribution of lipoprotein cholesterol classes. The Framingham study. Arteriosclerosis 3, 260–72 (1983).

36. Flynn, E. et al. Sex-specific genetic effects across biomarkers. Eur J Hum Genet 29, 154–163 (2021).

37. Zore, T., Palafox, M. & Reue, K. Sex differences in obesity, lipid metabolism, and inflammation-A role for the sex chromosomes? Mol Metab 15, 35–44 (2018).

38. AlSiraj, Y. et al. XX sex chromosome complement promotes atherosclerosis in mice. Nat Commun 10, 2631 (2019).

39. Natarajan, P. et al. Chromosome Xq23 is associated with lower atherogenic lipid concentrations and favorable cardiometabolic indices. Nat Commun 12, 2182 (2021).

40. Stacey, D. et al. ProGeM: a framework for the prioritization of candidate causal genes at molecular quantitative trait loci. Nucleic Acids Res 47, e3 (2019).

41. Zhu, X., Duren, Z. & Wong, W.H. Modeling regulatory network topology improves genome-wide analyses of complex human traits. Nat Commun 12, 2851 (2021).

42. Rondini, E.A., Duniec-Dmuchowski, Z., Cukovic, D., Dombkowski, A.A. & Kocarek, T.A. Differential Regulation of Gene Expression by Cholesterol Biosynthesis Inhibitors That Reduce (Pravastatin) or Enhance (Squalestatin 1) Nonsterol Isoprenoid Levels in Primary Cultured Mouse and Rat Hepatocytes. J Pharmacol Exp Ther 358, 216–29 (2016).

43. Wang, Y. et al. Therapeutic target database 2020: enriched resource for facilitating research and early development of targeted therapeutics. Nucleic Acids Res 48, D1031–D1041 (2020).

44. Ochoa, D. et al. Open Targets Platform: supporting systematic drug-target identification and prioritisation. Nucleic Acids Res 49, D1302–D1310 (2021).

45. Saiz-Vazquez, O., Puente-Martinez, A., Ubillos-Landa, S., Pacheco-Bonrostro, J. & Santabarbara, J. Cholesterol and Alzheimer’s Disease Risk: A Meta-Meta-Analysis. Brain Sci 10(2020).

46. Deb, S., Puthanveetil, P. & Sakharkar, P. A Population-Based Cross-Sectional Study of the Association between Liver Enzymes and Lipid Levels. Int J Hepatol 2018, 1286170 (2018).

47. Joshi, A.D. et al. Four Susceptibility Loci for Gallstone Disease Identified in a Meta-analysis of Genome-Wide Association Studies. Gastroenterology 151, 351–363 e28 (2016).

48. Ruth, K.S. et al. Using human genetics to understand the disease impacts of testosterone in men and women. Nat Med 26, 252–258 (2020).

49. Pirastu, N. et al. Genetic analyses identify widespread sex-differential participation bias. Nat Genet 53, 663–671 (2021).

50. Bhatt, D.K. et al. Hepatic Abundance and Activity of Androgen-and Drug-Metabolizing Enzyme UGT2B17 Are Associated with Genotype, Age, and Sex. Drug Metab Dispos 46, 888–896 (2018).

51. Nielsen, J.B. et al. Loss-of-function genomic variants highlight potential therapeutic targets for cardiovascular disease. Nat Commun 11, 6417 (2020).

52. Aragam, K.G. et al. Discovery and systematic characterization of risk variants and genes for coronary artery disease in over a million participants. medRxiv, 2021.05.24.21257377 (2021).

53. Votava, J.A. & Parks, B.W. Cross-species data integration to prioritize causal genes in lipid metabolism. Curr Opin Lipidol 32, 141–146 (2021).

54. Ahmad, N.N., Robinson, S., Kennedy-Martin, T., Poon, J.L. & Kan, H. Clinical outcomes associated with anti-obesity medications in real-world practice: A systematic literature review. Obes Rev, e13326 (2021).

55. Donaldson, J.G. & Jackson, C.L. Regulators and effectors of the ARF GTPases. Curr Opin Cell Biol 12, 475–82 (2000).

56. Marquer, C. et al. Arf6 controls retromer traffic and intracellular cholesterol distribution via a phosphoinositide-based mechanism. Nature Communications 7, 11919 (2016).

57. Hu, J. et al. Small GTPase Arf6 regulates diabetes-induced cholesterol accumulation in podocytes. Journal of Cellular Physiology 234, 23559–23570 (2019).

58. Sabatine, M.S. et al. Evolocumab and Clinical Outcomes in Patients with Cardiovascular Disease. New England Journal of Medicine 376, 1713–1722 (2017).

59. Schwartz, G.G. et al. Alirocumab and Cardiovascular Outcomes after Acute Coronary Syndrome. New England Journal of Medicine 379, 2097–2107 (2018).

60. Ray, K.K. et al. Two Phase 3 Trials of Inclisiran in Patients with Elevated LDL Cholesterol. New England Journal of Medicine 382, 1507–1519 (2020).

61. Nelson, C.P. et al. Genetic Assessment of Potential Long-Term On-Target Side Effects of PCSK9 (Proprotein Convertase Subtilisin/Kexin Type 9) Inhibitors. Circ Genom Precis Med 12, e002196 (2019).

62. Mahajan, A. et al. Fine-mapping type 2 diabetes loci to single-variant resolution using high-density imputation and islet-specific epigenome maps. Nat Genet 50, 1505–1513 (2018).

63. Assimes, T. et al. A large-scale multi-ethnic genome-wide association study of coronary artery disease. Nature Portfolio (2021).

64. Ridker, P.M. et al. Cardiovascular Efficacy and Safety of Bococizumab in High-Risk Patients. New England Journal of Medicine 376, 1527–1539 (2017).

65. Hopewell, J.C. et al. Differential effects of PCSK9 variants on risk of coronary disease and ischaemic stroke. Eur Heart J 39, 354–359 (2018).

66. Yu, X.H. et al. ABCG5/ABCG8 in cholesterol excretion and atherosclerosis. Clin Chim Acta 428, 82–8 (2014).

67. Silbernagel, G. et al. High intestinal cholesterol absorption is associated with cardiovascular disease and risk alleles in ABCG8 and ABO: evidence from the LURIC and YFS cohorts and from a meta-analysis. J Am Coll Cardiol 62, 291–9 (2013).

68. Teupser, D. et al. Genetic Regulation of Serum Phytosterol Levels and Risk of Coronary Artery Disease. Circulation: Cardiovascular Genetics 3, 331–339 (2010).

69. Artha, I. et al. High level of individual lipid profile and lipid ratio as a predictive marker of poor glycemic control in type-2 diabetes mellitus. Vasc Health Risk Manag 15, 149–157 (2019).

70. Hussain, A., Ali, I., Ijaz, M. & Rahim, A. Correlation between hemoglobin A1c and serum lipid profile in Afghani patients with type 2 diabetes: hemoglobin A1c prognosticates dyslipidemia. Ther Adv Endocrinol Metab 8, 51–57 (2017).

71. Chen, J. et al. The trans-ancestral genomic architecture of glycemic traits. Nat Genet 53, 840–860 (2021).

72. Chami, N. et al. Exome Genotyping Identifies Pleiotropic Variants Associated with Red Blood Cell Traits. Am J Hum Genet 99, 8–21 (2016).

73. Leong, A. et al. Mendelian Randomization Analysis of Hemoglobin A1c as a Risk Factor for Coronary Artery Disease. Diabetes Care 42, 1202–1208 (2019).

74. McDonagh, E.M. et al. PharmGKB summary: very important pharmacogene information for G6PD. Pharmacogenet Genomics 22, 219–28 (2012).

75. Dore, M.P., Parodi, G., Portoghese, M. & Pes, G.M. The Controversial Role of Glucose-6-Phosphate Dehydrogenase Deficiency on Cardiovascular Disease: A Narrative Review. Oxid Med Cell Longev 2021, 5529256 (2021).

76. Spielman, R.S. et al. Common genetic variants account for differences in gene expression among ethnic groups. Nat Genet 39, 226–31 (2007).

77. Zhu, A.Z. et al. Genetic and phenotypic variation in UGT2B17, a testosterone-metabolizing enzyme, is associated with BMI in males. Pharmacogenet Genomics 25, 263–9 (2015).

78. Yang, T.L. et al. Genome-wide copy-number-variation study identified a susceptibility gene, UGT2B17, for osteoporosis. Am J Hum Genet 83, 663–74 (2008).

79. Gencer, B. et al. Cardiovascular risk and testosterone - from subclinical atherosclerosis to lipoprotein function to heart failure. Rev Endocr Metab Disord 22, 257–274 (2021).

80. Firtser, S. et al. Relation of total and free testosterone and sex hormone-binding globulin with cardiovascular risk factors in men aged 24-45 years. The Cardiovascular Risk in Young Finns Study. Atherosclerosis 222, 257–62 (2012).

81. Schooling, C.M. et al. Genetic predictors of testosterone and their associations with cardiovascular disease and risk factors: A Mendelian randomization investigation. Int J Cardiol 267, 171–176 (2018).

82. Au Yeung, S.L. et al. Genetically predicted 17beta-estradiol and cardiovascular risk factors in women: a Mendelian randomization analysis using young women in Hong Kong and older women in the Guangzhou Biobank Cohort Study. Annals of Epidemiology 26, 171–175 (2016).

83. Winkler, T.W. et al. Quality control and conduct of genome-wide association meta-analyses. Nat Protoc 9, 1192–212 (2014).

84. Magi, R. et al. Trans-ethnic meta-regression of genome-wide association studies accounting for ancestry increases power for discovery and improves fine-mapping resolution. Hum Mol Genet 26, 3639–3650 (2017).

85. Willer, C.J., Li, Y. & Abecasis, G.R. METAL: fast and efficient meta-analysis of genomewide association scans. Bioinformatics 26, 2190–1 (2010).

86. Liu, D.J. et al. Meta-analysis of gene-level tests for rare variant association. Nat Genet 46, 200–4 (2014).

87. Winkler, T.W. et al. Approaches to detect genetic effects that differ between two strata in genome-wide meta-analyses: Recommendations based on a systematic evaluation. PLoS One 12, e0181038 (2017).

88. Giambartolomei, C. et al. Bayesian test for colocalisation between pairs of genetic association studies using summary statistics. PLoS Genet 10, e1004383 (2014).

89. Caliskan, M. et al. Genetic and Epigenetic Fine Mapping of Complex Trait Associated Loci in the Human Liver. Am J Hum Genet 105, 89–107 (2019).

90. Barbeira, A.N. et al. Exploring the phenotypic consequences of tissue specific gene expression variation inferred from GWAS summary statistics. Nat Commun 9, 1825 (2018).

91. Hindy, G. et al. Rare coding variants in 35 genes associate with circulating lipid levels – a multi-ancestry analysis of 170,000 exomes. bioRxiv, 2020.12.22.423783 (2021).

92. Brown, E.E. et al. Genetic testing in dyslipidemia: A scientific statement from the National Lipid Association. J Clin Lipidol 14, 398–413 (2020).

93. Hegele, R.A. et al. Rare dyslipidaemias, from phenotype to genotype to management: a European Atherosclerosis Society task force consensus statement. Lancet Diabetes Endocrinol 8, 50–67 (2020).

94. Zhu, X. & Stephens, M. Large-scale genome-wide enrichment analyses identify new trait-associated genes and pathways across 31 human phenotypes. Nat Commun 9, 4361 (2018).

95. Ge, T., Chen, C.Y., Ni, Y., Feng, Y.A. & Smoller, J.W. Polygenic prediction via Bayesian regression and continuous shrinkage priors. Nat Commun 10, 1776 (2019).

96. Purcell, S. et al. PLINK: a tool set for whole-genome association and population-based linkage analyses. Am J Hum Genet 81, 559–75 (2007).

97. Denny, J.C. et al. PheWAS: demonstrating the feasibility of a phenome-wide scan to discover gene-disease associations. Bioinformatics 26, 1205–10 (2010).

98. Verma, A. et al. A simulation study investigating power estimates in phenome-wide association studies. BMC Bioinformatics 19, 120 (2018).

99. Nelson, C.P. et al. Association analyses based on false discovery rate implicate new loci for coronary artery disease. Nat Genet 49, 1385–1391 (2017).

100. Liu, Z. et al. Causal relationships between NAFLD, T2D and obesity have implications for disease subphenotyping. J Hepatol 73, 263–276 (2020).

101. Zhou, W. et al. Efficiently controlling for case-control imbalance and sample relatedness in large-scale genetic association studies. Nat Genet 50, 1335–1341 (2018).

